# Revitalizing systemic immune responses in advanced NSCLC using FLT3L and SBRT

**DOI:** 10.1101/2025.01.29.25321332

**Authors:** Divij Mathew, Nitin Ohri, Shin Foong Ngiow, Balazs Halmos, KC Wumesh, Madhur Garg, Jeffrey M Levsky, Haiying Cheng, Rasim A. Gucalp, William Raymond Bodner, Rafi Kabarriti, Claudia Gutierrez-Chavez, Adel Zakharia, Shalom Kalnicki, Tibor Keler, Michael Yellin, Andy J Minn, Amit Maity, Josephine R Giles, Chandan Guha, E. John Wherry

**Affiliations:** Systems Pharmacology & Translational Therapeutics, University of Pennsylvania Perelman School of Medicine, Philadelphia, PA, USA; Department of Radiation Oncology, Montefiore Einstein Comprehensive Cancer Center, Bronx, NY, USA; Department of Oncology, Montefiore Einstein Comprehensive Cancer Center, Bronx, NY, USA; Department of Radiology, Montefiore Einstein Comprehensive Cancer Center, Bronx, NY, USA; Celldex Therapeutics, Hampton, NJ, USA; Parker Institute for Cancer Immunotherapy, University of Pennsylvania Perelman School of Medicine, Philadelphia, PA, USA; Abramson Family Cancer Research Institute, University of Pennsylvania Perelman School of Medicine, Philadelphia, PA, USA; Radiation Oncology, UPenn, Philadelphia, PA, USA; Mark Foundation Center for Immunotherapy, University of Pennsylvania Perelman School of Medicine, Philadelphia, PA, USA; Department of Radiation Oncology, University of Utah, Salt Lake City, Utah, USA; Institute for Immunology and Immune Health, University of Pennsylvania Perelman School of Medicine, Philadelphia, PA, USA

## Abstract

Non-small cell lung cancer (NSCLC) is the leading cause of cancer mortality worldwide. For patients who develop progressive disease after treatment with chemotherapy and immunotherapy, treatment options are limited. Here, we report the results of a Phase II clinical trial where twenty-nine patients with advanced, previously treated NSCLC were treated with stereotactic body radiotherapy (SBRT) targeting a single site of disease along with CDX-301, a recombinant human Fms-like tyrosine kinase 3 ligand (FLT3L). The primary study endpoint was progression-free survival four months after study entry (PFS4), which was achieved for 14 patients (48%). Abscopal responses were observed on fludeoxyglucose-18 positron emission tomography (FDG-PET) in nine patients (31%). The median overall survival for all participants was 18 months, with a median overall survival for participants with abscopal response of 31 months. To identify underlying immune signatures of response, we developed a high dimensional flow cytometric approach that quantified 31 distinct cell subsets from peripheral blood, and performed multiplex proteomic analysis of 92 proteins from plasma. SBRT combined with CDX-301 significantly increased circulating myeloid cells, including monocytes, myeloid derived suppressor cells (MDSCs) and dendritic cells (DCs). Among DCs, FceR1-expressing DC2 and DC3 subsets changed most robustly upon treatment. These dynamic changes in DCs and MDSCs following initiation of SBRT and CDX-301 returned to baseline by 8 weeks. However, these changes were also associated with treatment-induced T cell activation with quantitatively robust activation of CD4 T cells. Abscopal responses were associated with a prolonged increase in DC1 cells, Th1-like CD4 T cells, circulating IL-12 and FLT3L. Integrated analysis of multiple types of immune parameters revealed a strong association between coordinated DC induction, CD4 T cell and CD8 T cell activation, and inflammatory cytokine activity in patients with abscopal responses, compared to a stark lack of immune coordination in patients without abscopal responses. These coordinated immune responses were sustained for over 4 weeks in responders, highlighting a potential therapeutic axis engaged in a systemic immune response against multiple lesions in NSCLC. Overall, these findings underscore the potential of combining *in situ* vaccination, using SBRT, with strategies to enhance the activation of innate immune cells, such as DCs through FLT3L, to potentiate robust anti-tumor T cell responses.

## Introduction

NSCLC is the largest contributor to cancer-related fatalities worldwide. Effective options for advanced NSCLC may include platinum-doublet chemotherapy, immunotherapy targeting the PD-1/PD-L1 axis, and targeted therapy against oncogenic driver mutations. Patients for whom these options are exhausted have limited treatment options. Approved agents in this setting, which include docetaxel (+/- ramucirumab), gemcitabine, albumin-bound paclitaxel, and pemetrexed, yield response rates of only approximately 10%^1–3^. Novel treatment options for these patients are needed.

One of the mechanisms for clinical benefit of checkpoint blockade in NSCLC and other cancers is the reinvigoration of tumor-specific T cell responses. However, the lack of durable clinical responses to first-line treatment regimens including checkpoint blockade in NSCLC suggests T cell defects that are not sufficiently corrected by blocking the PD-1 pathway. For example, in addition to severe T cell exhaustion, lack of T cell priming, insufficient tumor antigen availability and/or the inability to induce new T cell responses following tumor mutation likely contribute to lack of durable clinical benefit in many NSCLC patients^4^. One strategy to increase tumor antigen availability and/or T cell priming is using stereotactic body radiotherapy (SBRT). This high-dose radiation induces immunogenic tumor cell death, releasing tumor antigens that can be taken up and presented by antigen-presenting cells. This process can potentially lead to the induction of new T cell responses or the more efficient restimulation of existing T cells^5^. Such effects occasionally manifest in abscopal responses in which even tumor lesions outside the radiation field regress following SBRT^6,7^. In mice, SBRT combined with PD-1 blockade can result in induction of tumor specific T cell responses^8^. Despite these data, the effectiveness of SBRT, even when combined with PD-1 blockade, remains suboptimal in humans and it remains challenging to induce effective abscopal responses following SBRT perhaps due to defects in innate immune cells and/or antigen presentation.

Fms-like tyrosine kinase 3 ligand (FLT3L) is a potent hematopoietic growth factor that mobilizes stem cells and greatly increases the number of circulating dendritic cells (DCs) in blood and tissues. FLT3L can initiate immune responses and cause tumor regression in preclinical models of fibrosarcoma^9^, breast cancer^10^, prostate cancer^11^, and lung adenocarcinoma^12^, however FLT3L has not been demonstrated to have clinical activity when given as a single agent in clinical trials. Using a mouse model of lung adenocarcinoma, we previously found that FLT3L treatment alone partially reduced tumor burden and had increased efficacy when combined with high-dose radiotherapy^12^. This effect from combined treatment was lost when T cells were ablated. Because FLT3, the receptor for FLT3L is mainly expressed on hematopoietic and myeloid lineage cells, these data suggested involvement from both myeloid and lymphoid compartment in controlling disease^12^. An early Phase I clinical trial reported the therapeutic application of FLT3L in patients with metastatic colon cancer and NSCLC. FLT3L was used to expand DCs *in vivo* for isolation and antigen loading *in vitro* with peptides derived from carcinoembryonic antigen (CEA). Immunization with these CEA-loaded DCs resulted in expansion of antigen-specific T cells^13^. Furthermore, a Phase II clinical trial in melanoma patients tested CDX-301 in combination with poly-ICLC, a Toll-like receptor 3 agonist, and anti-DEC205-NY-ESO-1, to target NY-ESO-1 peptide to antigen presenting cells. This combination resulted in induction of NY-ESO-1 specific T cell responses. Although this study was not designed to evaluate clinical benefit, the induction of antigen specific T cells highlights the potential of pairing FLT3L with tumor-derived antigens for induction of adaptive immune responses^14^.

Given these previous results, we hypothesized that the release of tumor antigen from SBRT in conjunction with expanded DCs from FLT3L would be able to stimulate effective anti-tumor immune responses despite progression after previous treatments. To test this idea, we performed a Phase II clinical trial (NCT02839265) in NSCLC combining treatment with a recombinant human FLT3L (CDX-301) with SBRT in patients who had progressed after at least one line of standard systemic therapy. The combination of CDX-301 and SBRT resulted in significant clinical effects, including abscopal responses, which indicate a systemic immune response capable of tumor regression beyond the irradiated lesion. This treatment increased the frequency of circulating dendritic cells (DCs), myeloid-derived suppressor cells (MDSCs), and monocytes. Notably, CD4 T cell activation and expansion were more robust than CD8 T cells. The induction of various DC subsets and correlations between innate and adaptive immune responses distinguished patients with and without abscopal benefits. Abscopal responses were associated with a coordinated Type 1 immune response, marked by increases in DC1s, Th1-like CD4 T cells, and circulating IL-12. Overall, the CDX-301 and SBRT treatment showed promising clinical activity in patients with progressing NSCLC, with clinical benefits linked to a sustained and coordinated Type 1 immune response in those with abscopal responses. This suggests the potential to harness the vaccinal effects of SBRT by enhancing adaptive immune responses through the induction or rejuvenation of antigen-presenting cells, including DCs.

## Results

### FLT3L and SBRT achieves clinical endpoint in recurrent metastatic NSCLC

To investigate the clinical efficacy of CDX-301 and SBRT in NSCLC patients who progressed on previous therapies, we conducted a Phase II clinical trial. Twenty-nine patients with advanced NSCLC were enrolled between October 2016 and January 2020 (**Fig. 1A, SFig 1A**). All patients had previously received at least one line of systemic therapy for advanced NSCLC, and most had been treated with multiple previous lines of therapy (median 3 types of treatment, range 1 to 5). Twenty-six patients (90%) had previously been treated with immune checkpoint inhibitors targeting the PD-1/PD-L1 axis, with a median interval of 4 months from termination of immune checkpoint inhibitor therapy to study registration (**SFig 1B**). Twenty-two of those patients (85%) received immunotherapy as second-line therapy or beyond. Twelve patients (41%) had performance status (PS) 2 at study entry. Overall, patients had a median of four hypermetabolic lesions (range 2 to 12) on baseline PET/CT at the time of study entry.

**Figure 1:**
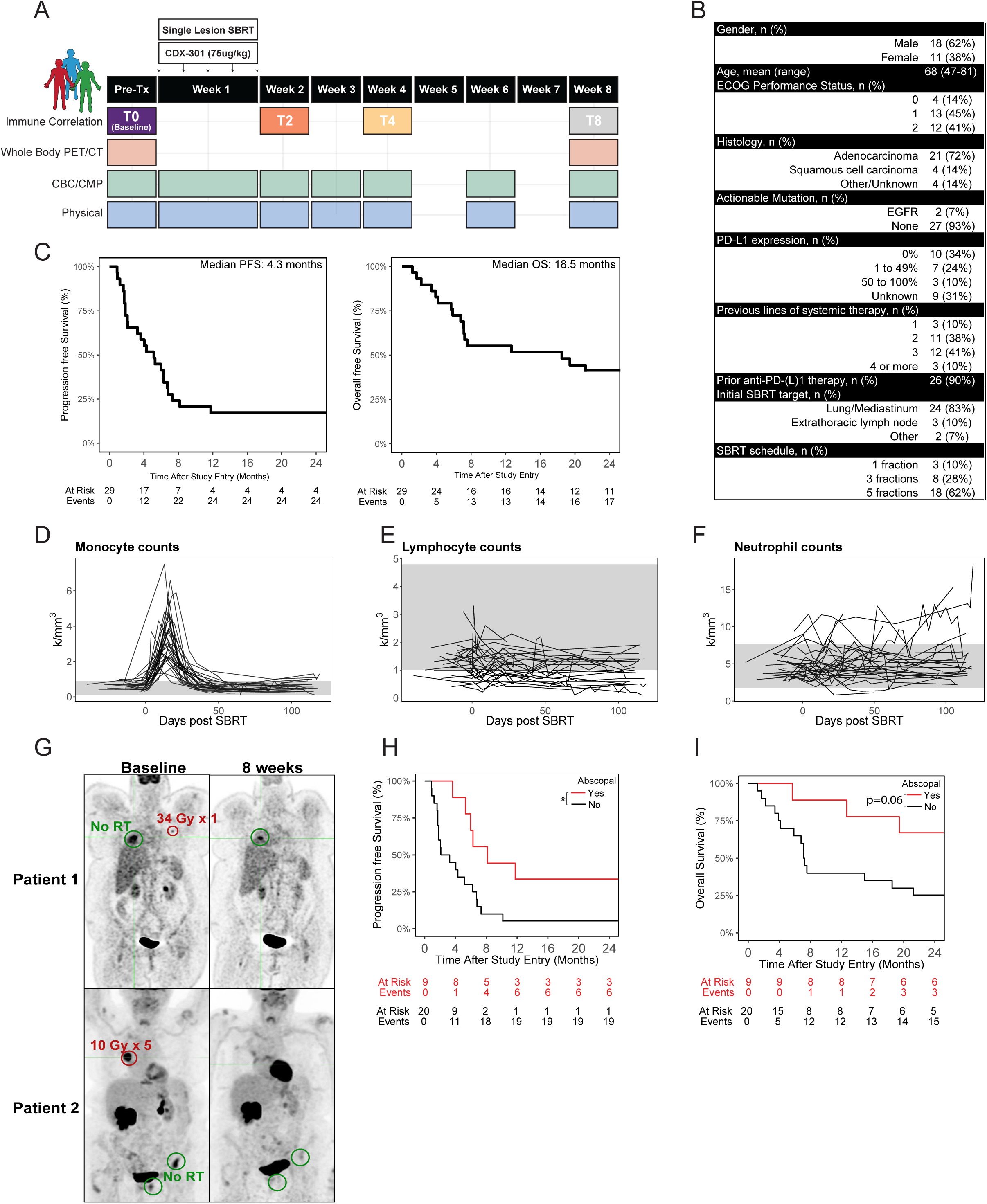
FLT3L and SBRT achieves clinical endpoints in recurrent metastatic NSCLC. (A) Study Schema for NCT02839265 with different modalities collected at listed timepoints. (B) Clinical chart of 29 patients enrolled in study (C-D) Kaplan Meyer curves describing PFS (left) and OS (right) with median times listed all patients in the study. Cell counts of monocytes (D), lymphocytes (E), or neutrophils (F) after treatment initiation. Heathy donor range is shaded in grey. (G) PET imaging demonstrating abscopal responses for two study patients with index lesion (green) and irritated lesion (red) marked. (h-i) Kaplan-Meier curves of PFS (H) and OS (I) for patient subgroups with and without abscopal responses. P value determined by Log rank test. * p<0.05

All patients received 5 daily subcutaneous injections of CDX-301 (75 µg/kg) concurrent with SBRT (30-54 Gy in 1-5 fractions based on target size and location) directed at a single site of disease. For 23 participants (79%), the initial SBRT target was a lung tumor or thoracic lymph node. Fourteen patients (48%) achieved a progression-free survival (PFS) of at least four months, satisfying our prespecified efficacy endpoint. Nine patients (31%) later received a second 1-week cycle of SBRT and CDX-301. The median time interval between the first and second treatment cycles was 6 months (range: 4 to 26 months), and no patient received other cancer therapy between treatment cycles (**Fig.1B**). The median PFS duration following initial study treatment was 4.3 months, and the median overall survival (OS) duration was 18.5 months (**Fig.1C**). Exploratory analyses of prognostic factors revealed that 2-4 lesions at study entry was significantly associated with PFS and OS, consistent with observations that immunotherapy has greater efficacy with reduced tumor burden^15^ (**SFig 1C**). Blood counts showed an increase in monocytes after treatment with little change in neutrophil or lymphocyte count (**Fig.1D-Fig.1F**), consistent with FLT3L inducing monocyte differentiation from hematopoietic stem cells (HSC). No grade 4-5 adverse events related to study therapy were observed. One participant experienced grade 3 gastrointestinal symptoms possibly related to study therapy. Despite including patients with poor performance status (PS2), no patient discontinued study therapy early due to treatment-related toxicity, demonstrating a favorable toxicity profile for this treatment Notably, in 31% of the patients, regression of disease outside the irradiated field was observed on PET imaging, indicating an abscopal response (**Fig. 1G, SFig. 1D**). This result suggested that SBRT and CDX-301 combination treatment induces a systemic immune response capable of anti-tumor activity in some patients. PFS was improved in patients exhibiting abscopal responses, with a trend towards improved overall survival (OS) as well (**Fig. 1H-1I**). There were no significant differences in age, prior anti-PD(L)1 therapy, or histology between response groups (**SFig 1E**). Thus, the combination of CDX-301 and SBRT induced abscopal responses in a subset of patients suggesting the induction of systemic immune responses even in this heavily pretreated NSCLC patient population.

### Expansion of DC2 and DC3 post FLT3L and SBRT identifies abscopal responses

The increased monocyte numbers in the clinical blood counts suggested that treatment with CDX-301 and SBRT altered the immune cell composition. To understand global immune system alterations after treatment, we designed a 26-parameter high dimensional flow cytometry panel to interrogate the major leukocyte populations in the peripheral blood (**SFig. 2A**). Among CD45+ cells, CDX-301 and SBRT treatment was associated with increased frequencies of DCs (increased 135.6% compared to baseline), MDSCs (increased 148.9%), and monocytes (increased 23.3%) 2 weeks post treatment initiation (**SFig. 2B**). The frequency of DCs remained elevated through 4 weeks post treatment, and changes for all cell types returned to baseline by 8 weeks (**SFig. 2B-C**). To examine DC populations in more detail, we used expression of CD141+ and FceR1a to identify DC1 and DC2 that have been associated with Type 1 and Type 2 responses respectively^16^. Recent studies have identified additional heterogeneity within the DC2 compartment based on expression of CD163, CD5, and CD14 ^17,18^ that allowed us to further subdivide these DCs into DC2 and DC3 subsets (**Fig. 2A**). DC2 and DC3 robustly expanded in the blood of most patients following treatment whereas changes in the DC1 subset were quantitatively more modest and more variable between patients. These alterations returned to baseline by 8 weeks (**Fig. 2B**). Within the DC2 and DC3 subsets, CD5+ DC2, CD5-DC2, and CD14+ DC3 expanded at 2 weeks post treatment. There were negligible changes in the frequencies of preDCs and CD14+ DC3 and a transient reduction in pDCs at 2 weeks post treatment (**Fig. 2C, SFig. 2D**). To test which DC subsets changed the most throughout treatment, we used a repeated measure ANOVA test. Indeed, CD5-DC2, CD14-DC3, and CD5+DC2 had the most significant changes associated with time among the DC subsets in response to CDX-301 and SBRT (**Fig. 2D**, **Fig. 2E**).

**Figure 2:**
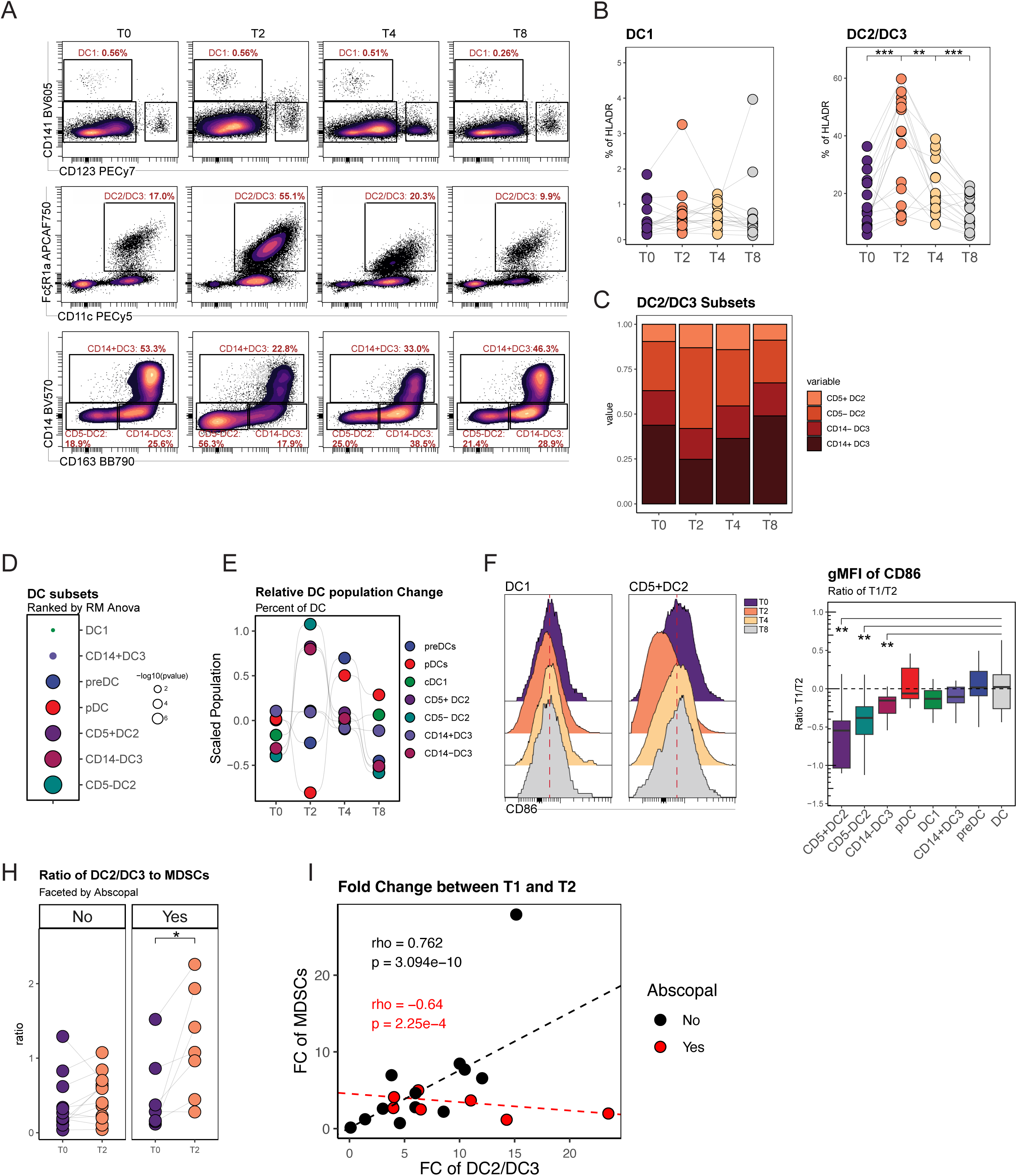
Expansion of DC2 & DC3 after FLTL and SBRT is associated with Abscopal Effects. (A) Gating scheme to identify DC1 (top row), DC2/DC3 (middle row), and subsets of DC2/DC3 (bottom row) after treatment at T0 (B) Paired analysis of 18 patients at 4 timepoints measuring changes in DC1 (left) and DC2/DC3s (right). Significance was determined using a paired Wilcox test with ns p>0.05, ** p<=0.01, *** p<=0.001. (C) Stacked bar plots over 4 timepoints reflecting percentage change of CD5+DC2, CD5-DC2, CD14-DC3, and CD14+DC3 in the DC2/DC3 gate. (D) DC subsets ranked in ascending order of p value determined by repeated measure ANOVA with time as the independent variable and the different DC subsets as dependent variables. Outlines indicate p <0.05. (E) Summary bump plot of scaled DC populations over time. (F) Histogram of CD86 expression in DC1(left) and CD5+DC2 (right) changing with time. (G) Ratio of the gMFI of CD86 of different DC subsets at T0 to T2 compared to the total DC population. (H) Ratio of % DC2/DC3 to % MDSC faceted by clinical Abscopal response. Significance was determined using a paired Wilcox test with p <= 0.05. (I) Scatter plot with patient coordinates represent by FC of DC1/DC2 and MDSC from T2 to T1 colored by Abscopal response. Rho and p values determined by Spear-man’s rank correlation with non-parametric trend lines (Theil-Sen)

Therapeutic administration of FLT3L can induce HSC mobilization and differentiation of bone marrow derived DCs^19^. To test if the increase in DC populations in the blood after CDX-301 and SBRT was due to de novo DC differentiation from the bone marrow or expansion of DCs in the periphery, we examined CD86 as a proxy of DC maturation^20^. A significant reduction in CD86 expression was observed on CD5+DC2, CD5-DC2, and CD14-DC3 2 weeks after treatment indicating an immature differentiation state (**Fig. 2F**, **Fig. 2G**). This result suggests that the treatment primarily stimulated *de novo* differentiation of immature DC subsets from the bone marrow (**Fig. 2F**, **Fig. 2G**).

In addition to stimulating DC differentiation, FLT3L can generate myeloid-derived suppressor cells^21^ (MDSCs). MDSCs have immune-suppressive effects that can contribute to tumor progression. Indeed, CDX-301 and SBRT treatment induced a significant increase in CD14+ monocyte-like cells in the blood, consistent with MDSC-like cells (**SFig. 2C**, **2E**). To begin to interrogate how increases in both anti-tumor and pro-tumor myeloid populations related to clinical responses, we examined the ratio of these populations (DC2+DC3 to MDSCs) at baseline (T0) and Timepoint 2 (T2). Patients with an abscopal response had a significantly increased DC2+DC3:MDSC ratio whereas no such change was apparent in patients lacking an abscopal response (**Fig. 2H**). Furthermore, patients with an abscopal response had minimal change in the MDSC-like population between T1 and T2, in contrast to the increase in DC2s and DC3s following treatment (**Fig. 2I**). Thus, whereas most patients had an increase in DC2 and DC3 populations after treatment, abscopal responders were distinguished by the relative absence of change in MDSC-like cells in the blood at two weeks after treatment initiation.

### Prolonged CD4+ T cell responses after CDX-301 and SBRT are associated with abscopal responses

We next investigated whether these changes in the myeloid compartment were associated with altered T cell activation or differentiation. We developed a 26-parameter cytometry panel to analyze T cell activity over time (**SFig. 3A**). First, we asked whether SBRT and CDX-301 treatment resulted in T cell activation. Co-expression of HLA-DR, a marker of recent activation together with the cell cycle maker Ki67 identified robust increases in the frequency of activated CD4 T cells and, to a lesser extent, CD8 T cells at 2 weeks post therapy (**Fig. 3A-B, SFig 3B**). These responses of CD4 T cells in most patients were sustained at 4 weeks before returning to baseline by 8 weeks (**Fig. 3A-B**), consistent with previous reports of strong CD4 T cell responses after CDX-301^14^.

**Figure 3:**
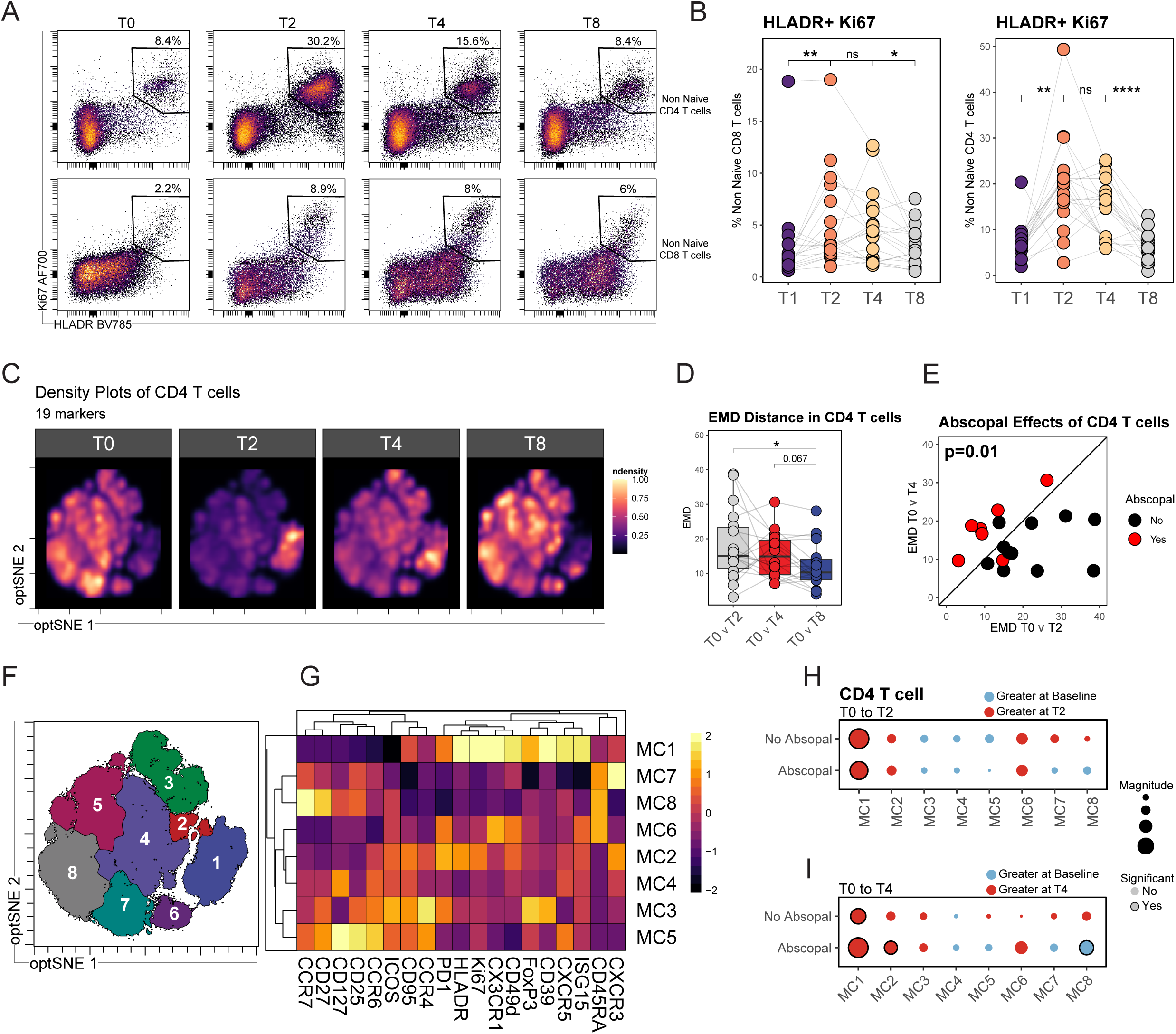
FLT3L and SBRT drives Th1 like CD4 T cell responses in Abscopal Responders. (A) Representative flow plots of activated (HLADR+Ki67+) CD4 non naïve T cells (top row) or activated CD8 non naïve T cells (bottom row) (B) Paired analysis of 18 patients at 4 times points measuring HLADR+KI67+ in non-naïve CD8 T (left) or non-naïve CD4 T (right). Significance was determined using a paired Wilcox test with ns p>0.05, * p<0.05, ** p<=0.01, *** p<=0.001, **** p<=0.0001. (C) Density plots of 500K CD4 T cells equally sampled by timepoint. (D) Earth mover distance between patient optSNE projections of CD4 T cells at indicated timepoints. Significance was determined using a paired Wilcox test with * p <0.05. (E) Scatter plot of EMD distance between T0 – T2 (x axis) and T0 – T4 (y axis) with black line representing a slope of 1 and color representing Abscopal response. P value was determined by a two-sample test for proportions. (F) FlowSOM clustering of 500K CD4 T cells broken into 8 meta clusters. (G) Heatmap highlighting the gMFI marker expression for each meta cluster scaled to column. (H-I) Dot plots of each meta cluster faceted by Abscopal response and separated by comparison of baseline to T2 (H) and baseline to T4 (I). Color of the dot indicates which timepoint has greater expression of the meta cluster, the size of the dot indicates the magnitude of the expression, and the outline represent if the expression is significant. Significance is a paired Wilcox test with outline representing p<0.05.

To further investigate changes in T cell activation and differentiation following treatment, we projected CD4 and CD8 T cell flow cytometry data into unique optSNE spaces and assessed changes over time (**Fig. 3C, SFig. 4A**). Density plot projections of the CD4 T cell data revealed a strong shift in overall phenotype at 2 weeks that returned to baseline by 8 weeks. Earth Mover’s Distance (EMD), a measurement of the minimum cost required to transform one distribution into another^22^, was used to quantify the magnitude of change between each time point. As expected, the greatest EMD value, indicating the largest effect on change in CD4 T cell differentiation, occurred from baseline to 2 weeks with the smallest effect from baseline to 8 weeks, (**Fig. 3D**). EMD values from baseline to two weeks and baseline to 4 weeks were similar, suggesting changes in the CD4 T cell population were sustained through 4 weeks post treatment. We next asked whether there were differences in amount of change to CD4 T cell differentiation in patients who experienced clinical abscopal responses compared to those who did not. Thus, we used the baseline:T2 and baseline:T4 EMD values as cartesian coordinates and projected each patient as a unique point on a scatter plot (**Fig. 3E**). Indeed, patients with an abscopal response had a greater EMD ratio at 4 weeks, whereas patients who did not experience an abscopal response had a greater EMD ratio at 2 weeks (**Fig. 3E**). These observations suggested an association of delayed and/or sustained changes to CD4 T cell activation and differentiation with abscopal anti-tumor responses.

**Figure 4:**
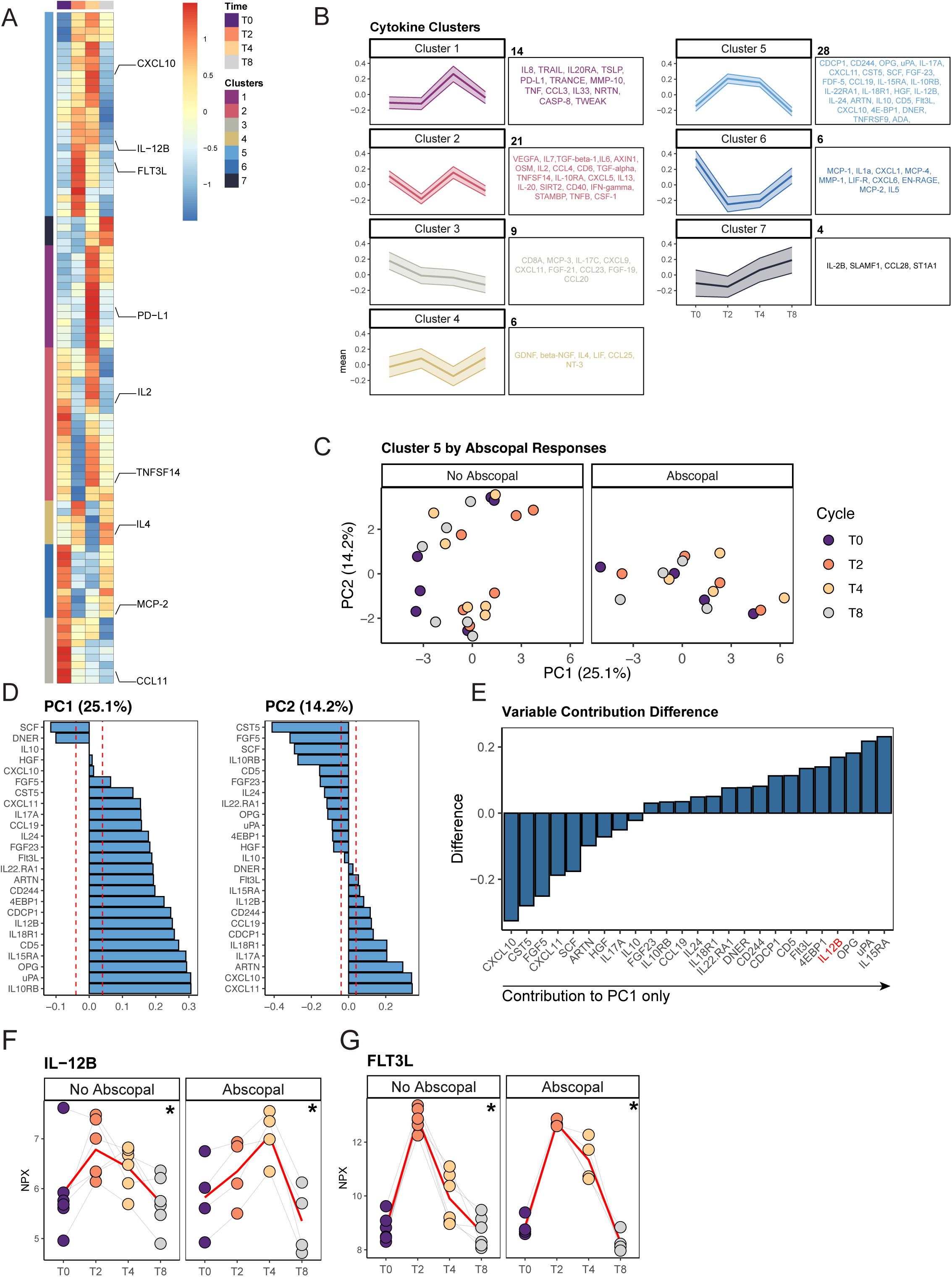
Abscopal responders have delayed upregulation of IL21 4 weeks post treatment. (A) Patient level PCA using (A) Heatmap of the mean of the NPX unit of 10 patients across each cytokine at each timepoint with 7 clusters identified by color. Clusters were determined by hierarchical clustering. (B) Temporal kinetics of cytokine patterns faceted by cluster. List and number of cytokines are listed adjacent to cluster temporal kinetics. (C) PCA of all cluster 5 cytokines colored by timepoint and faceted by Abscopal response. (D) PCA input parameters ranking contribution to positive or negative PC1 direction (left) and PC2 direction (right). Parameters within the redline indicate contributions to PC axis below equal distribution. (E) Variable contribution of PC1 subtract by the variables contribution in PC2. High values represent variables contributing just to PC1 movement. NPX expression of IL12B (F) or FLT3L (G) over time colored by timepoint. P value determined by repeated measure ANOVA with *p<0.05.

To investigate what aspects of altered CD4 T cell differentiation were responsible for these EMD changes, we applied FlowSOM clustering focusing on comparing patients with and without abscopal responses. This unbiased analysis identified eight CD4 T cell meta clusters (MC1-8) (**Fig. 3F**, **Fig. 3G**). Both patient response groups had significant increases in MC1 at two weeks after treatment (**Fig. 3H**). MC1 contained activated HLA-DR^hi^ Ki67^hi^CX3CR1^hi^ CD39^hi^ PD1^hi^ CD4 T cells (**Fig. 3H**. CD39 can also be expressed by regulatory CD4 T cells (Tregs)^23^ and FoxP3 was also increased in MC1. However, even after excluding FoxP3+ CD4 T cells, there was still a substantial increase in CD45RA^Lo^CD27^−^ CD4 T cells expressing CX3CR1^24^ and CD39, consistent with MC1 (**SFig. 3D, SFig. 3E**). We next asked if this population was unique to CDX-301 and SBRT treatment or was also found in response to other therapies. Indeed, a population of CX3CR1+CD39+ CD4 T cells could be identified in the blood of NSCLC patients treated with anti-PD1 monotherapy (**SFig. 3F**). However, there was no change in the frequency of this population during anti-PD-1 therapy (**SFig. 3F**) suggesting that, although these cells were normally found in the blood of NSCLC patients, the expansion of this activated CD4 T cell population was associated with the combination CDX-301 and SBRT treatment. Taken together, treatment with CDX-301 and SBRT generated a highly activated CD4 population in abscopal and non-abscopal patients within two weeks of initiation of therapy.

Given the changes in CD4 T cell responses in abscopal responders at 4 weeks, we hypothesized that differences in the CD4 T cell differentiation state between clinical response groups would diverge at this timepoint. To investigate this possibility, we calculated the fold change of all MCs from baseline to T4. MC2 and MC8 were significantly different in abscopal responders, but not in patients who did not experience an abscopal effect (**Fig. 3I**). MC8, consistent with naïve CD4 T cells (CD45RA+CD27+CCR7+), was lower in abscopal patients at T4. In contrast, MC2, an activated Th1-like subset (CXCR3+PD-1+HLA-DR+Ki67+), was increased at week 4. Both patient groups also had an increase in MC1 at T4 versus baseline, consistent with the T2 changes. However, MC2, increased only in abscopal patients, had lower FoxP3, CD39 and CX3CR1 compared to MC1 but increased PD-1, ICOS, CXCR3 and CCR6 (**Fig. 3I**). Notably, ICOS+ activated CD4 T cells have been implicated in effective responses to CTLA-4 blockade^25^, another setting that may require or induce new CD4 T cell priming. Thus, these data suggested that CDX-301 and SBRT treatment may be associated with increases in Th1-like activated CD4 T cells. Moreover, this change in highly activated Type 1 CD4 T cell responses was only sustained in patients who experienced an abscopal response.

CDX-301 and SBRT treatment also resulted in increased CD8 T cell activation. However, the overall magnitude of changes in the CD8 T cell responses were more modest than for the CD4 T cells and quantification using EMD did not reveal any statistically significant changes in density distribution over time (**SFig. 4A-B**). Projection of EMD values from baseline to T2 and baseline to T4 as cartesian coordinates also did not separate abscopal response patients from non-abscopal response patients based on CD8 T cell responses (**SFig. 4C**). To further investigate changes in the CD8 T cell responses following treatment, we used FlowSOM and identified 5 MC. Two clusters, MC1 and MC2 increased in response to therapy (**SFig. 4D-F**). MC1 had features of exhausted like CD8 T cells with high expression of PD1 and CD39, but also expressed Ki67 and HLA-DR suggesting possible response to therapy and/or recent reinvigoration as has been observed in settings of PD-1 blockade^26,27^. MC2 shared features of activated effector CD8 T cells with high Ki67 and HLA-DR, but also expression of CD45RA, CX3CR1 and ISG15. This MC2 cluster had some similarities to terminally differentiated effector memory RA cells (i.e. CD8 T cells re-expressing CD45RA). Indeed, manual gating confirmed an increase in CD45RA+ CD27-TEMRA-like CD8 T cells following treatment as well as an increase in PD-1+CD39+ exhausted-like CD8 T cells (**SFig. 4F-H**). Thus, CDX301 and SBRT treatment resulted in activation of both CD4 and CD8 T cells. However, the response induced from CD4 T cells was greater in magnitude than for CD8 T cells and this CD4 T cell response was characterized by Th1-like cells. Moreover, sustained CD4 T cell responses were associated with abscopal responses.

### Induction of circulating IL-12 in abscopal responders at 4 weeks post treatment

By week 4 after treatment, patients with abscopal responses had significantly more Th1-like CD4 T cells. Th1-like CD4 T cell responses are associated with Type 1 polarizing inflammatory cytokines, including interleukin-12 (IL-12)^28^. To test whether abscopal patients displayed a Th1 polarizing cytokine response to treatment compared to non-abscopal patients, we examined changes in 92 cytokines in the serum. To understand how cytokines changed over time, we hierarchically clustered all cytokines on a heatmap revealing 7 temporal patterns (**Fig. 4A-B**). Cluster 5 had the greatest increase from baseline to T2, capturing cytokines induced in response to CDX-301 and SBRT treatment, whereas cluster 6 identified cytokines decreased in response to treatment. Repeated measures ANOVA ranked the cytokines most changed in abscopal responders and non-abscopal responders over 8 weeks (**SFig. 5A**). This analysis revealed FLT3L as the top cytokine, consistent with CDX-301 treatment. In addition, the top changed cytokines in both clinical response groups were enriched for those contained in cluster 5, confirming that cytokines in this cluster were both responsive to treatment and the most altered over time (**SFig. 5A**). To further investigate which of the 28 cytokines from cluster 5 contributed most to changes observed after treatment in each patient group, we performed principal component (PC) analysis (**Fig. 4C**). Patients with abscopal responses were primarily characterized by variation in PC1, whereas non-abscopal responders had substantially more contribution of PC2 and less variance in PC1 (**Fig. 4C**). To understand this distinct cytokine pattern in abscopal responders, we examined how each cluster 5 cytokine contributed to the PC1 and PC2 axis (**Fig. 4D**). We focused on cytokines that predominantly influenced the direction of PC1 while exerting lesser influence on PC2 because of the strong association of abscopal response patients with PC1, but not PC2. Among these cytokines, interleukin 12B (IL-12B) emerged as one of the top contributors (**Fig. 4D-E**). The kinetics of IL-12B was distinct between patients with and without abscopal responses, continuing to rise from baseline to T4 in abscopal patients, whereas the peak of IL-12B was at T2 in non-abscopal patients (**Fig. 4F**). This pattern of IL-12B was mirrored by the Th1-like CD4 T cell MC in abscopal patients described above. In preclinical models, *IL12p40* mRNA increases in the spleen after FLT3L infusion^29^. To test whether a relationship existed between FLT3L and IL-12 in the blood in NSCLC patients, we examined the kinetics of FLT3L in both abscopal response groups. As expected, there was a similar increase in FLT3L in both patient groups from T0 to T2 likely representing administered CDX-301. However, the subsequent decline from this peak at T2 was considerably more robust in non-abscopal patients whereas abscopal patients had sustained FLT3L protein detectable in blood (**Fig. 4G**). This sustained FLT3L could reflect differences in CDX-301 drug pharmacodynamic and/or induction of host FLT3L. Nevertheless, this difference could explain the sustained IL-12B observed in blood and may underlie the sustained Type 1 cellular responses identified in the T cell analysis above.

**Figure 5:**
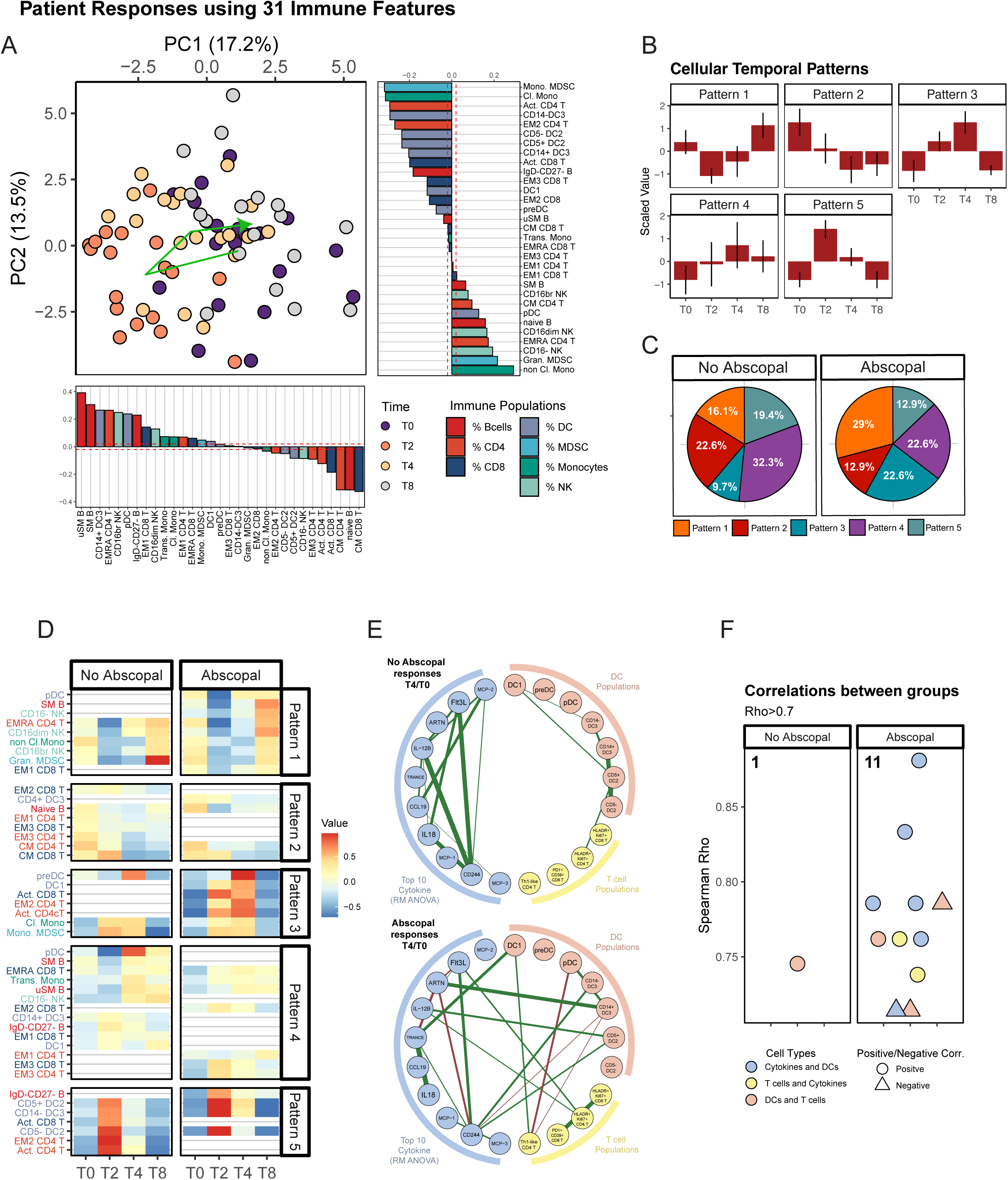
Systems integration of cellular responses of SBRT and FLT3L. (A) Patient level PCA using aggregated flow cytometry data representing 31 unique immune populations. Each point represents a patient colored by timepoint with the green arrow representing the mean of PC1 and PC2 at each timepoint. Variable contributions for the PC1 axis (right) and PC2 axis(bottom) are ranking by magnitude and direction of contribution and colored by parent population. (B) Identified temporal cellular patterns identified from the 31 immune subsets changing over time. (C) Composition of the temporal cellular patterns in non-Abscopal patients and Abscopal patients (D) Cellular composition of each temporal pattern faceted by Abscopal response. (E) Spearman rank correlations of DC1, preDC, pDC, CD14-DC3, CD14+DC3, CD5+DC2, CD5-DC2, Th1-like CD4 T cells (CXCR3+), exhausted CD8 T cells (PD1+CD39+), activated CD4 T cells (HLADR+Ki67+), activated CD8 T cells (HLADR+Ki67+), and the top 10 cytokines from that are changing the most with time. Nodes are colored by associated group (cytokine, T cells, and DCs). Green edges represent positive correlations and red edges represent negative correlations. Correlations >0.7 for inclusion in correlation network. (F) Dot plot of spearman correlations that are >0.7 between nodes of different groups and faceted by abscopal responses.

### Abscopal responders induce Type1 immunity in response to CDX301 and SBRT

To further investigate the distinct Type 1 CD4 T cell and cytokine responses identified in abscopal responders we next performed a comprehensive integrated analysis of leukocytes populations in the blood following CDX-301 and SBRT treatment. We built a patient level PC plot using 31 unique immune subsets from the two high dimensional flow cytometry panels described above. To test how the immune responses changed over time, we overlayed a vector (indicated by the arrow in **Fig. 5A**) that intersected the mean of PC1 and PC2 at each time point. The circular trajectory of the arrow suggested that treatment induced a transient change in overall immune composition that returned towards baseline by 8 weeks (**Fig. 5A**). To test which PC axis best explained the treatment effect, we examined the PC1 and PC2 coordinates over the 4 timepoints. PC1 significantly changed between each timepoint, whereas PC2 changed substantially only from week 2 to week 4, suggesting the PC1 axis likely explains the immunological variance in response to SBRT and CDX-301 between abscopal and non-abscopal patients (**SFig. 6A-B**). To determine which immune population(s) contributed to this response to treatment, we plotted the cellular contributions to the PC1 and PC2 axes. The initial shift along the PC1 axis to the left is largely explained by activated CD4 T cells, DC2 and DC3 subsets, consistent with the observations above. However, changes in monocytic MDSC-like cells and classical monocytes had a greater contribution to this change in PC1 to the left than conventional DCs or CD4 T cells, suggesting an effect of FL3L driving increases in monocytes as well as DCs (**Fig. 5A**). To test if time from PD1/PDL1 inhibition altered responses to CDX-301 and SBRT, we calculated the fold change from baseline to T2, T4 or T8 of each leukocyte population and correlated those changes to time from last PD-1-based immunotherapy treatment (**SFig. 6C-D**). These data suggested that more recent treatment with a PD-1 pathway antagonist was associated with greater change in DC1 cells from T1 to T4. In contrast longer duration since last anti-PD-1/PD-L1 correlated with larger fold change in CM CD8 T cells and CD15+ MDSCs (**SFig. 6C-D**). Although these observations suggest that recent treatment with immune checkpoint blockade could have an influence on combined CDX-301 and SBRT therapy that is worth further exploration, the major Th1-like CD4 T cell response could not be explained by residual PD-1 or PD-L1 blocking antibodies due to recent treatment.

A subset of patients received a second round of CDX-301 and SBRT treatment (**SFig. 7A**). We were, therefore, able to use this smaller group of patients to test whether additional treatment resulted in a similar immune response. Although fewer patients were available limiting statistical power, the overall trajectory in PC space was consistent with that observed following initial treatment. Indeed, the T0 and T2 coordinates between the two cycles of treatment was consistent, supporting the notion that the second round of CDX-301 and SBRT stimulated a similar pattern of immune response as the initial treatment cycle (**SFig. 7B-C)**. For example, activated CD4 and CD8 T cells increased after a second cycle of treatment with again a trend to greater magnitude of a CD4 T cell response (**SFig. 7D**). Thus, CDX-301 and SBRT treatment is capable of repeatedly inducing innate and adaptive immune changes in NSCLC patients.

The observations above revealed a transient change in the immune system after treatment with SBRT and CDX-301. Next, we investigated whether immune response trajectory differed between abscopal and non-abscopal patients. First, we used the 31 immune cell subsets and identified 5 temporal patterns (**Fig. 5B**). We then compared these patterns between non-abscopal and abscopal patients (**Fig. 5C**). Indeed, abscopal response patients displayed a distinct overall temporal pattern compared to non-abscopal patients. Pattern 3 exhibited the greatest change between the groups with > two-fold increase in abscopal responders compared to non-abscopal responders. To identify which immune population contributed to each temporal group, we plotted immune cell subpopulation for abscopal versus non-abscopal patients. Pattern 3 for abscopal patients captured the increase in activated CD4+ T cells, activated CD8+ T cells, and DC1s from baseline to T4 and the return to baseline by T8. In non-abscopal patients, however, these immune cell types were found in Pattern 4 and Pattern 5 reflecting more transient and/or lower magnitude of change (**Fig. 5D**). Indeed, whereas manual gating of all patients did not reveal a clear pattern in the DC1 response (**Fig. 2B**), when examined by abscopal response, there was a clear increase in DC1s from T0 through T4 in abscopal but not non-abscopal patients (**SFig. 8A**). To further investigate the kinetics of changes in DC populations and activated T cells, we scaled the different leukocyte populations such that each cell type exists on the same dynamic range, allowing comparison of temporal changes. CD5-DC2, CD5+ DC3, and CD14-DC3 peaked at T2 in both abscopal and non-abscopal patients before returning to baseline by T8 (**SFig. 8B**). In non-abscopal patients activated CD4 and CD8 T cells also peaked at T2 returning to baseline thereafter. However, in abscopal patients activated CD4 T cells and DC1 initially increased at T2, but then continued to increase and peaked at T4 before returning to baseline (**SFig. 8C**), consistent with the kinetics of Temporal Pattern 4 identified above.

We next tested if the patterns observed for DCs and T cells also occurred in an independent cohort of NSCLC patients treated with anti-PD1 monotherapy^30^. However, despite immune activation following anti-PD-1 treatment, the immune populations induced by anti-PD-1 had a distinct pattern from that observed following CDX-301 and SBRT (**SFig. 8B,8C**). For example, anti-PD-1 monotherapy provoked a continued increase in CD8 T cell activation with a decline in DC1 populations (**SFig. 8D**), while abscopal responders displayed a proportional increase in DC1 compared to both non abscopal patients and patients treated with anti-PD1 monotherapy (**SFig. 8E**). These observations suggested that the combination of SBRT and CDX-301 induces distinct immune dynamics and coordinated signature that is not observed in anti-PD1 monotherapy and further separates non-abscopal responders and abscopal responders.

The distinct temporal patterns of immune response observed, in patients with versus without abscopal responses suggested potential differences in synchronization of responses of different immune populations. Thus, we next investigated whether sustained coordination of individual immune responses might distinguish patient groups. To test this idea, we calculated the fold change from T0 to T4 of all 7 DC populations (DC1, preDC, pDC, CD14-DC3, CD14+DC3, CD5+DC2, CD5-DC2), 4 T cells populations (Th1-like CD4 T cells, exhausted CD8 T cells, activated CD4 T cells, activated CD8 T cells), and the 10 cytokines that changed most robustly throughout treatment. We built a network correlation plot separated by abscopal response. In the absence of an abscopal response, few interactions were observed among DCs, T cells, and cytokines (**Fig. 5E**). In contrast, for abscopal responders, a highly interconnected network of 11 correlations between distinct immune cell types and cytokines was identified, with a notable correlation between DC1 and Th1-like CD4 T cells (**Fig. 5E, 5F**). Thus, a key difference in response to CDX-301 and SBRT in patients who experienced systemic immune responses defined by clinical abscopal response was the induction of well-coordinated immune responses temporally connecting different features of innate and adaptive immunity. These findings highlight the ability of this treatment combination to induce a sustained Type I immune response, characterized by the presence of IL12, DC1, and Th1-like CD4 T cells associated with abscopal anti-tumor responses in a subset of patients.

## Discussion

These studies highlight the immunological effects of combining tumor destruction using targeted radiation and innate immune cell induction through FLT3L treatment. This combination of treatment provoked a coordinated innate and adaptive immune response in a subset of previously treated NSCLC patients that was associated with an abscopal anti-tumor response and clinical benefit. Previous studies have used FLT3L to expand DCs and concurrent administration of a known tumor antigen^13,14^ leading to the generation of antigen-specific T cell responses. Clinically, however, many tumors continue to evolve antigenically^31^ and, if driver antigens are not targeted, these tumors can escape T cell responses targeting a single or limited number of epitopes. One approach to overcome antigenic tumor evolution is through *in situ* vaccination approaches such as SBRT. Indeed, use of single lesion ablative SBRT can induce immunogenic death in tumor cells providing an opportunity for T cell priming and/or restimulation^32^ to a variety of tumor epitopes. In preclinical models, FLT3L alone failed to improve survival of mice with breast cancer that were treated with sub-ablative SBRT doses (8Gy x 3), commonly used in clinical trials of SBRT and immune checkpoint blockade^33^. However, ensuring effective induction of T cell responses also requires competent innate immunity including effective antigen presenting cells such as DC. Use of FLT3L to induce and expand DC is one potential approach to engage effective DC induction during other therapies^12^. Indeed, in preclinical models, ablative doses of SBRT synergizes with FLT3L in reducing tumor burden and extending survival in mice^34^, but it has been unclear how this combinational therapy will extend to humans. Thus, the data presented here demonstrate that combination of FLT3L and ablative SBRT directed at a single malignant lesion was well tolerated, induced abscopal responses, and achieved PFS endpoints revealing immunological and potentially clinical opportunities with this combination.

As immunotherapy now plays a role in the management of numerous malignancies, the possibility that radiotherapy could augment the efficacy of immunotherapy has garnered considerable interest ^35,36^. Additionally, the synergistic use of immunotherapy and SBRT markedly elevates abscopal response rates (ARR), which are notably minimal when either modality is administered in isolation^8^. Moreover, several studies have pointed to induction of T cell responses in settings of abscopal response^32,37^ provoking the hypothesis that immunogenic cell death through radiation primes (or boosts) T cell responses. These observations also suggest, however, that, although adding checkpoint blockade increases the likelihood of abscopal effect, removing T cell inhibition is unlikely to be the only barrier to improving the immunological effectiveness of SBRT. An additional challenge in many tumors is the absence of sufficient immunostimulatory APC including DC. FLT3L is a potent stimulator of DC responses and previous studies using FLT3L and peptide vaccination in cancer have demonstrated strong induction of T cell responses^38,39^. Here, our data document induction of DCs following FLT3L treatment of NSCLC patients. Futhermore, the specific subsets, kinetics and coordination of DC induction with other immune responses were notable. DC are heterogenous and have been subdivided based on marker expression and functionality^40,41^. Human DC1 are efficient stimulators of CD8 T cell responses but can also generate CD4 T cell responses against tumor antigen *in vivo*^42^. DC2 are thought to be potent stimulators of CD4 T cells^43^. DC3 can be distinguished from DC2 by expression of CD163 and may also stimulate CD8 T cells^44^. pDC are potential producers of IFN and unlikely to play a major role in induction of T cell responses through antigen presentation directly. Several other populations of DC, including mregDC (that may have some similarity to DC3) also have been described^45^. In previously treated NSCLC patients FLT3L treatment induced robust quantitative changes in DC2/3 populations and analysis of maturation markers suggested these were newly generated cells. Although numerically less abundant, DC1s were also increased compared to baseline. pDC were also detectable in the blood, but their temporal induction lagged the DC1 and DC2/3 populations. Thus, FLT3L treatment in previously treated NSCLC patients was capable of robust induction of multiple DC populations when used in combination with SBRT. Although maturation markers suggest many of the DC in the blood are newly derived from bone marrow precursors, it is also possible that some of the relevant DC are mobilized from the tumor tissue by FLT3L. Moreover, whether these changes in circulating DC populations reflect changes in tumor tissue or tumor draining lymph nodes remains to be determined.

The changes in DC populations following CDX-301 and SBRT were also temporally associated with induction of T cell responses. In animal models, an abscopal anti-tumor effect requires T cells^46^ and both preclinical models and human observations support the importance of dynamic changes in T cell responses following SBRT combined with different immune-based therapies. In many cancer immunotherapy settings, CD8 T cells are thought to be the key mediators of antitumor activity. Indeed, PD-1 pathway blockade is associated with “reinvigoration” of CD8 T cell responses leading to control of disease in many types of cancer^26^. The importance of CD4 T cells in effective tumor immunity has long been appreciated^47^ and recent work highlights the potential role for “triads” of CD4 and CD8 T cells together with DC in tumor tissue where CD4 T cells enable a more effective CD8 T cell response^48^. Other studies have also begun to revise the concept that CD4 T cells only provide help to CD8 T cells. Indeed, CD4 T cells can lead to tumor control even in the absence of CD8 T cells, an effect that is lost in the absence of Tbet or CD11c+ DCs^49^. In fact, Th1-like CD4+ T cells can eliminate tumors that escape CD8+ T cell control because of loss of MHC^50^. The combination of CDX-301 and SBRT here provoked both a CD8 T cell response, and an even more quantitatively robust CD4 T cell response in the blood. It was notable that, although CD8 T cells can be potent anti-tumor effectors in many cases, the patients with systemic benefit evidenced by abscopal responses here were those with the most robust induction of in Th1+ like CD4 T cells. An intriguing possibility is that, in addition to the clear effects of FLT3L on DC populations, this cytokine may act directly on CD4 T cells, perhaps to limit IL-10 and augment CD4 T cell activation^51^. The relative importance of FLT3L directly on CD4 T cells versus through effects on DC populations in this current setting remains unclear. Moreover, although in the current studies it was not possible to determine which T cells were tumor-specific because of the lack of tumor tissue for TCR comparison, these data suggest a key role for CD4 T cell responses in this setting.

One of the more remarkable observations in this study was the distinct coordination of immune responses in abscopal patients compared to the absence of coordination in non-abscopal patients. By integrating the analysis of temporal changes in key immune cell types and circulating inflammatory mediators in the blood following CDX-301 and SBRT treatment, clearly distinct patterns of interaction were apparent between innate immune cells (DC subset), circulating cytokines, and dynamically changing T cell populations. One cytokine of particular importance for induction of Th1 responses is IL-12, which can be produced by DC^52,53^. Indeed, in agreement with the generation of DC responses by FLT3L treatment, IL-12 increased in the serum in both abscopal and non-abscopal patients. However, in abscopal patients this IL-12 response persisted longer and the temporal extension of this cytokine response was also associated with a prolonged response of DC1, activated CD4 and activated CD8 T cells. Moreover, an integrated analysis of different components of the immune response over time demonstrated coordinated responses between circulating inflammatory mediators, DC subsets, and T cell responses supporting the notion that induction of systemic anti-tumor immunity in abscopal patients reflects a multi-layered immune response. Indeed, the evidence of coordination of effective DC responses with potent pro-immunity cytokines like IL-12 in these abscopal patients may point to future opportunities to enhance responses in those patients who do not achieve clinical benefit.

Thus, these studies highlight the combination of CDX-301 and SBRT as an approach to reactivate the immune system in NSCLC who have previously progressed on other therapies. Moreover, the sustained Type 1 immune response in abscopal responders and excellent safety profile indicate a therapeutic axis to that could be further harnessed to achieve systemic immunity against metastatic lesions. Given the favorable tolerability CDX-301 with SBRT, further exploration is warranted for administrating CDX-301 at later timepoints to perpetuate the systemic Type 1 response and anti-tumor activity. Finally, these data might also point to immune features lacking in patients who do not achieve clinical benefit and highlight a possible role for further augmenting antigen presentation and induction of innate immunity and overall coordination of key mediators of innate and adaptive immunity in settings such as SBRT intended to enhance tumor cell death and adaptive immune priming.

## Acknowledgement

We thank patients, families, and health care providers who participated in this study. Thanks to B. Degregorio and D.M. Anderson for infrastructure support and the Guha lab for support and discussions which led to the completion of this study. We thank past and present members of the Wherry and Guha laboratories for insightful discussions and laboratory support including Maura Jones, Amy Baxter and Brady Galan.

## Code Availability

### Funding

National Institutes of Health grant T32 CA009140 (JRG)

Cancer Research Institute-Mark Foundation Fellowship (JRG)

National Institutes of Health grant AI155577 (EJW)

National Institutes of Health grant AI115712 (EJW)

National Institutes of Health grant AI117950 (EJW)

National Institutes of Health grant AI108545 (EJW)

National Institutes of Health grant AI082630 (EJW)

National Institutes of Health grant CA210944 (EJW)

Mark Foundation for Cancer Research (EJW)

Parker Institute for Cancer Immunotherapy (EJW, DM)

### Author Contributions

Conceptualization: NO, TK, MY, CG

Methodology: DM, NO

Investigation: DM, NO, BH, MG, JML, HC, RAG, WRB, RK, AZ, SK, CGC

Formal Analysis: DM, NO, SFN, WK, AJM

Visualization: DM, NO

Writing – original draft: DM, JRG

Writing – review & editing: DM, NO, BH, JRG, EJW, CG

Funding Acquisition: CG and EJW

Supervision: CG and EJW

### Competing Interests

EJW is a member of the Parker Institute for Cancer Immunotherapy which funds cancer immunology research in his lab. EJW is an advisor for Arsenal Biosciences, Coherus, Danger Bio, IpiNovyx, New Limit, Marengo, Pluto Immunotherapeutics, Related Sciences, Santa Ana Bio, and Synthekine. EJW is a founder of and holds stock in Coherus, Danger Bio, and Arsenal Biosciences.

AJM. has received research funding from Merck. He has also received honoraria from Merck, AstraZeneca, Pfizer, and Takeda. AJM is an inventor on patents filed or issued on the IFN pathway and modified CAR T cells. He is a scientific founder for Dispatch Biotherapeutics and advisor for Related Sciences, Diagonal Therapeutics, and Xilio

JRG is a consultant for Arsenal Biosciences and Cellanome.

TK is an employee and stockholder in Celldex Therapeutics, Inc.

MY was an employee at Celldex Therapeutics, Inc. and is a stockholder.

BH is a consultant for Astra Zeneca, Boehringer Ingelheim, Apollomics, Janssen, Takeda, Merck, BMS, Genentech, Pfizer, Eli-Lilly, Arcus, Merus, Daiichi, Precede, BMS and has received research funding from Boehringer Ingelheim, Astra Zeneca, Merck, BMS, Advaxis, Amgen, AbbVie, Daiichi, Pfizer, GSK, Beigene, Janssen, Black Diamond Therapeutics, Forward Pharma, Numab, Arrivent.

## Methods

### Study Patients

Patients with histologically proven NSCLC who were previously treated with at least one line of standard systemic therapy for advanced disease were eligible for this phase II trial (NCT02839265). Other key eligibility criteria included ECOG performance status 0-2, radiological assessment within 21 days of study registration demonstrating at least one tumor or metastasis that would be amenable to SBRT as well as at least one other measurable site of disease. Patients with a history of brain metastases treated with local therapy and without evidence of progressive disease in the brain were eligible.

### Study Therapy

Subcutaneous CDX-301 (75 µg/kg) was administered daily for five days, starting on a Monday. During the same week, patients were treated with SBRT targeting a single hypermetabolic lung tumor, regional lymph node, or metastasis. SBRT targets were chosen by the treating physicians, primarily based on safety considerations. SBRT schedules were based on target location and followed NCCN guidelines and cooperative group trial recommendations. Dosimetric constraints for nearby structures were adopted from the same sources. Study patients received no other cancer therapy until disease progression occurred. Additional “cycles” of SBRT (to distinct lesions) and CDX-301 could be administered starting four months after initial study therapy, at the discretion of the treating physicians.

### Evaluations

Clinic evaluations, including toxicity evaluation using CTCAE v. 4.0, were performed prior to treatment, during weeks 1, 2, 3, 4, 6, and 8, and every 8 weeks thereafter. PET/CT was prior to treatment and every 8 weeks after treatment. Plasma for correlative studies was drawn prior to treatment, during weeks 2, 4, and 8, and every 8 weeks thereafter.

Radiographic responses were primary scored using immune-related response criteria (irRC), in which reduction of the sum of the products of the two largest perpendicular diameters (SPD) of at least 50% qualifies as a partial response (PR), and SPD increase of at least 25% from the nadir can qualify as progressive disease (PD) if confirmed on a second scan(12). Responses were also assessed on PET using volumetric PERCIST criteria, where a reduction in total glycolytic activity (TGA) of at least 45% qualifies as a partial metabolic response (PMR), and TGA increase of at least 75% qualifies as progressive metabolic disease (PMD)(13). Of note, SBRT target lesions were excluded from radiographic response scoring.

### Adverse Events

Study treatment well tolerated, with no treatment-related grade 3 or higher adverse events (AEs) except for one subject who experienced grade 3 gastrointestinal symptoms (abdominal pain, nausea, vomiting, and diarrhea) that were considered to possibly be related to treatment. The most common grade 1-2 adverse events that were possibly related to study therapy were fatigue (38%), dyspnea (24%), vomiting (21%), decreased appetite (17%), and nausea (17%). No patient discontinued study therapy due to treatment-related toxicity.

### Study Endpoints and Statistical Analyses

Patient characteristics, treatment details, toxicity data, and response rates were reported using descriptive statistics. The primary study endpoint was progression-free survival 4 months after initiation of study therapy (PFS4), scored using irRC(12). We hypothesized that PFS4 would be 20% or less with standard care (null hypothesis), and PFS4 would be improved to 40% or greater with study therapy (alternative hypothesis). We would deem the study regimen worthy of further study if PFS4 were achieved in 10 out of 29 study patients, yielding >80% probability of pursuing this regimen for further study if the alternative hypothesis were true and <5% probability of pursing this regimen if the null hypothesis were true.

Progression-free survival (PFS) and overall survival (OS) rates after initiation of study therapy were calculated using the Kaplan-Meier method. Median follow-up duration following study entry was calculated using the “reverse Kaplan-Meier” method(14). Cox proportional hazards models and logrank tests were used to assess potential prognostic factors in exploratory analyses.

### Antibody panels and staining

Approximately 1 × 10^6^ to 5 × 10^6^ were thawed from in RPMI media containing 1x DNase and 10% FBS. PBMCs were stained with live/dead mix (100μl, 10 min, RT), with FC block. Samples were washed with fluorescence-activated cell sorting (FACS) buffer, and spun down (1600 rpm, 5 min, RT). With the T cell panel, pellets was resuspended in 25 μl of chemokine receptor staining mix and incubated at 37°C for 20 min. After incubation, 25 μl of surface receptor staining mix was directly added, and the PBMCs were incubated at RT for a further 45 min. For the myeloid panel, 50uL of surface receptor antibody was directly added and PBMCs were kept at RT for 45min. After RT incubation, PBMCs from both panels were washed (FACS buffer, 1600 rpm, 5 min, RT) and resuspended in 50 μl of secondary antibody mix for 20 min at RT and then washed again (FACS buffer, 1600 rpm, 5 min, RT). Samples were fixed and permeabilized by incubating in 100 μl of Fix/Perm buffer (RT, 30 min) and washed in Perm Buffer (1800 rpm, 5 min, RT). PBMCs were stained with 50 μl of intracellular mix from their respect panels and kept overnight at 4°C. The following morning, samples were washed (Perm Buffer, 1800 rpm, 5 min, RT) and resuspended in PBS. For the surface receptor and chemokine staining mix, antibodies were diluted in FACS buffer with 50% BD Brilliant Buffer (BD, catalog no. 566349). Intracellular mix was diluted in Perm Buffer. See table for buffer information and table for antibody panel information.

### Flow cytometry

Samples were acquired on a five-laser BD FACS Symphony A5. Standardized SPHERO rainbow beads (Spherotech, catalog no. RFP-30-5A) were used to track and adjust photomultiplier tubes over time. UltraComp eBeads (ThermoFisher, catalog no. 01-2222-42) were used for compensation.

### Olink

Cytokines were measured from heparin-plasma using the Olink Extension Assay (PEA) to measure 92 unique analytes. Oligonucleotide labeled capture antibodies bind target cytokines and subsequently hybridize. The hybridized product is amplified and measured by qPCR, allowing for parallel detection of 92 cytokines within the same sample.

## Data availability

The CSV files containing the exported counts and serum proteomics data will be made available upon publication.

## Code availability

No custom code was developed in the analysis of this study.

---

**SFigure 1.**
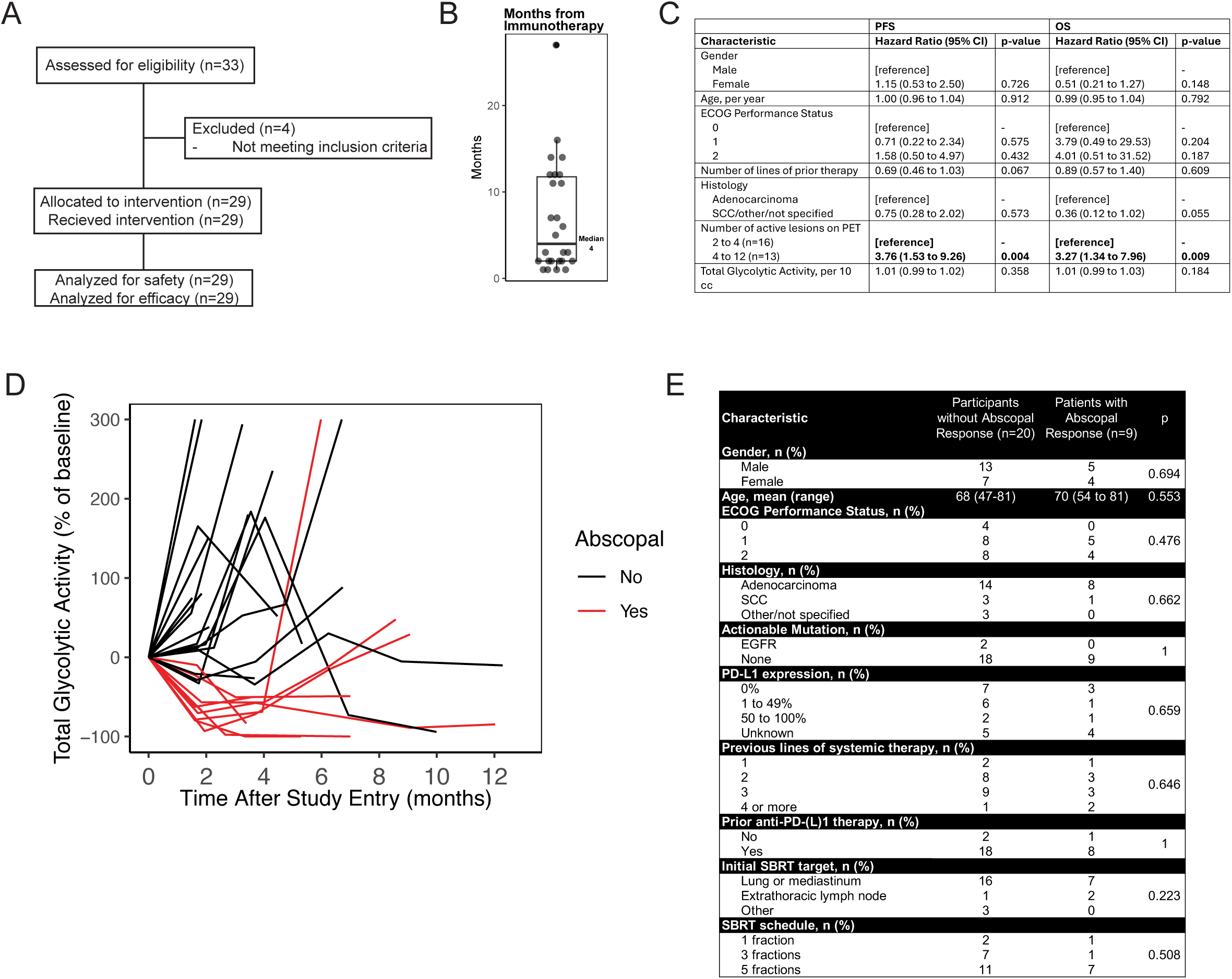
(A) CONSORT flow diagram for NCT02839265 of combining stereotactic body radiotherapy (SBRT) with FLT3 ligand immunotherapy (B) Time after immunotherapy of all patients enrolled in study with median time listed. (C) Cox model predicting characteristics that predict response to treatment. (D) Spider plots colored by Abscopal responses. (E) Clinical chart of patients enrolled in study broken into Abscopal responses and no Abscopal responses.

**SFigure 2.**
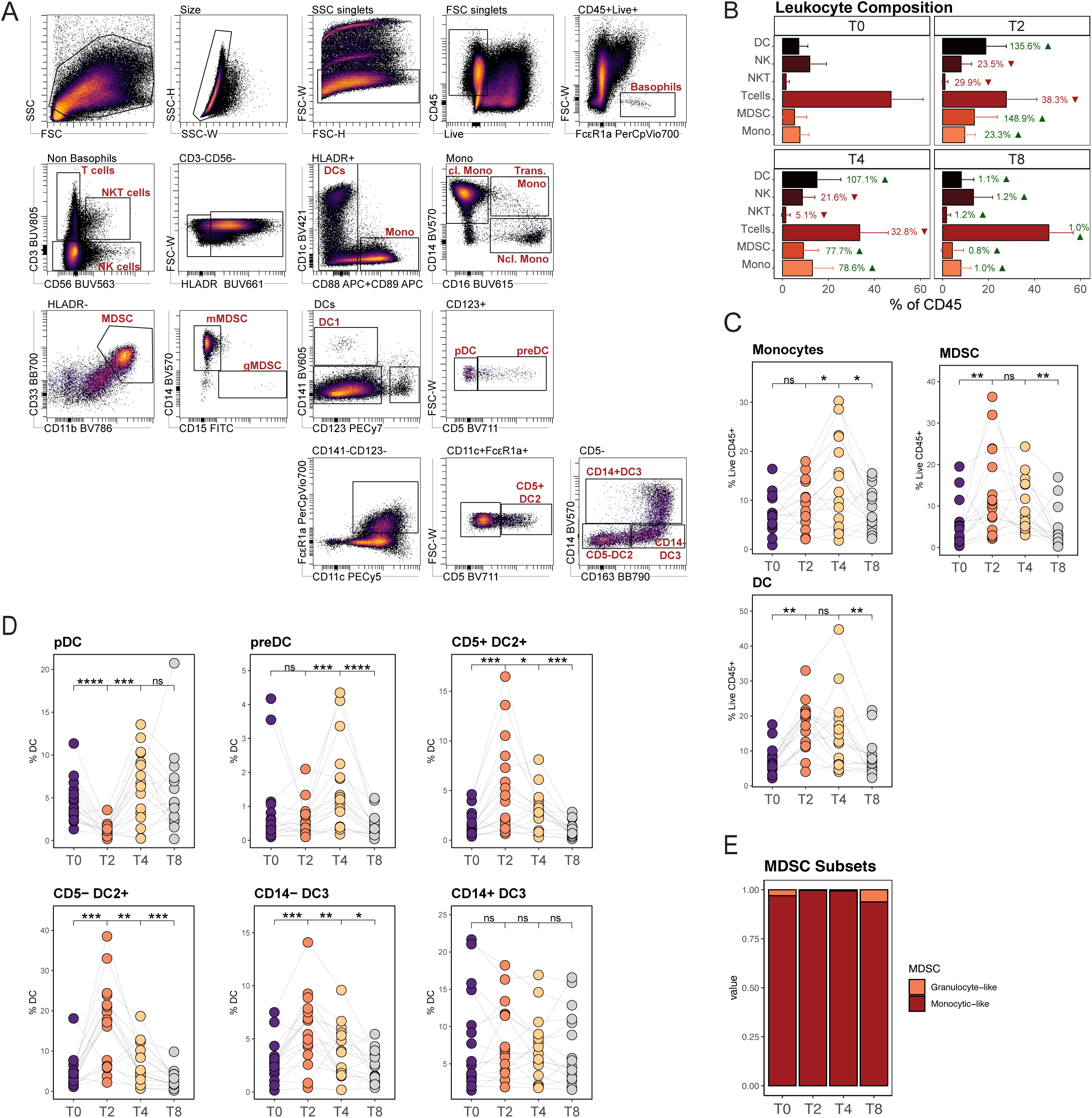
(A) Flow cytometry gating scheme to identify myeloid cell populations. (B) Leukocyte composition over time relative to T0. Green arrow and green colored percentage represents an increase from T0 and red arrow and red color percentage represents a decrease from T0. (C) Changes of Monocytes, MDSCs, and DC are a percent of total live CD45+ cells and colored by timepoint. (D) Changes of pDC, preDC, CD5+DC2+, CD5-DC2+, CD14-DC3, and DC14+DC3 as a percent of HLADR+ cells. (E) Stacked bar plots of change in granulocytic (CD15+) and monocytic (CD14+) MDSCs over time

**SFigure 3.**
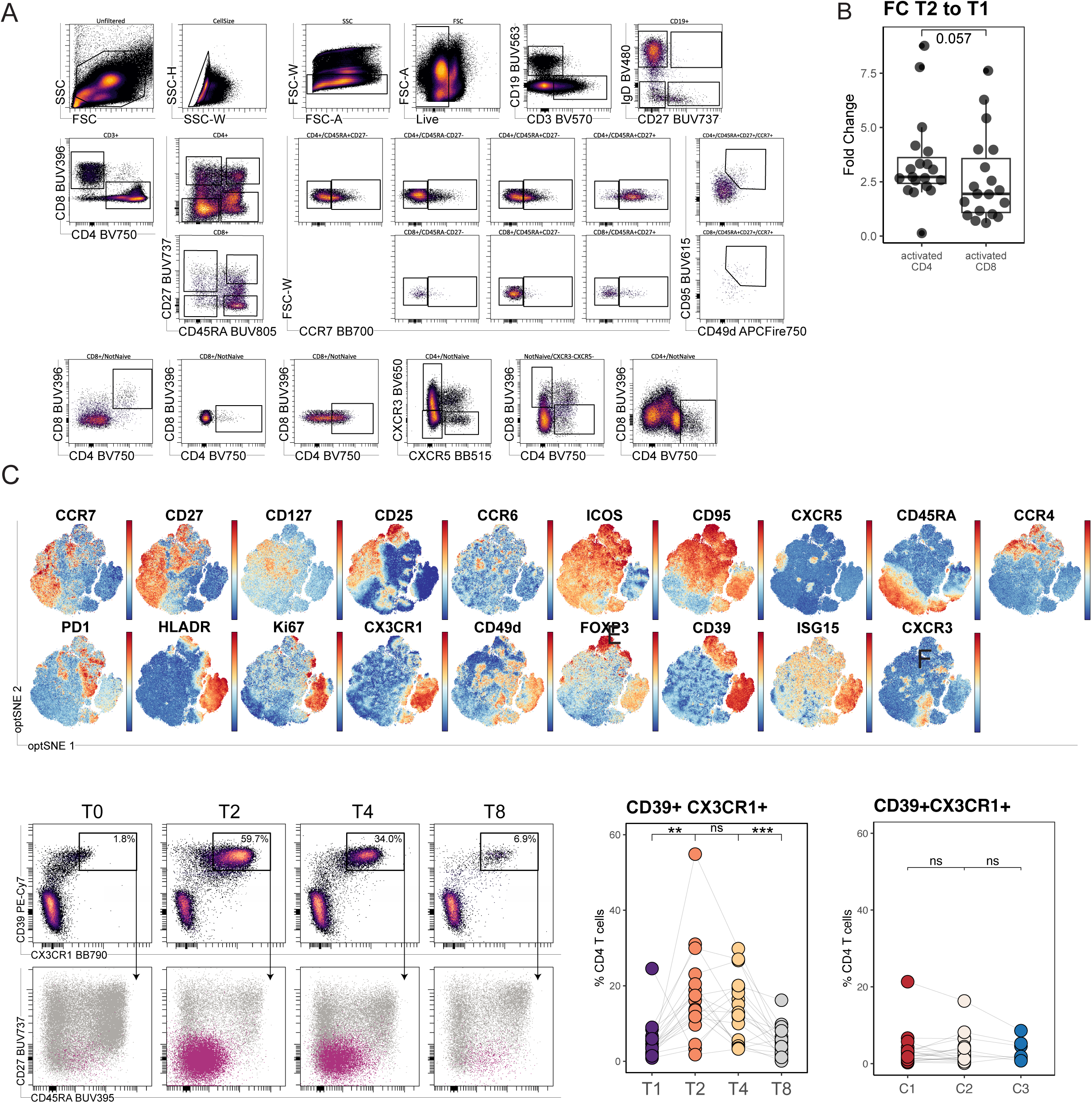
(A) Flow cytometry gating scheme to identify lymphocyte subsets. (B) Comparison of the fold change of CD4 and CD8 T cell activation from T2 to T1. Listed p value was determined uses a Wilcox test. (C) Feature plot of different protein markers which shape the CD4 optSNE embedding. (D) Biaxial flow plot depicting the expression of CX3CR1 and CD39 in FoxP3- CD4 T cells over time (top). Biaxial plot of CD45RA and CD27 with total CD4 T cells (grey) and CD39+CX3CR1+ (purple) shown (bottom). (E) Paired analysis of 18 patients at 4 times points measuring CX3CR1+CXCR3+ in non naïve CD4 T cells. Significance was determined using a paired Wilcox test with ns p>0.05, ** p<=0.01, *** p<=0.001. (F) Paired analysis of 23 patients over 3 cycles measuring CX3CR1+CXCR3+ in non naïve CD4 T cells. Patients were from a different trial (NCT03425006) using only pembrolizumab monotherapy. Significance was determined using a paired Wilcox test with ns p>0.05.

**SFigure 4.**
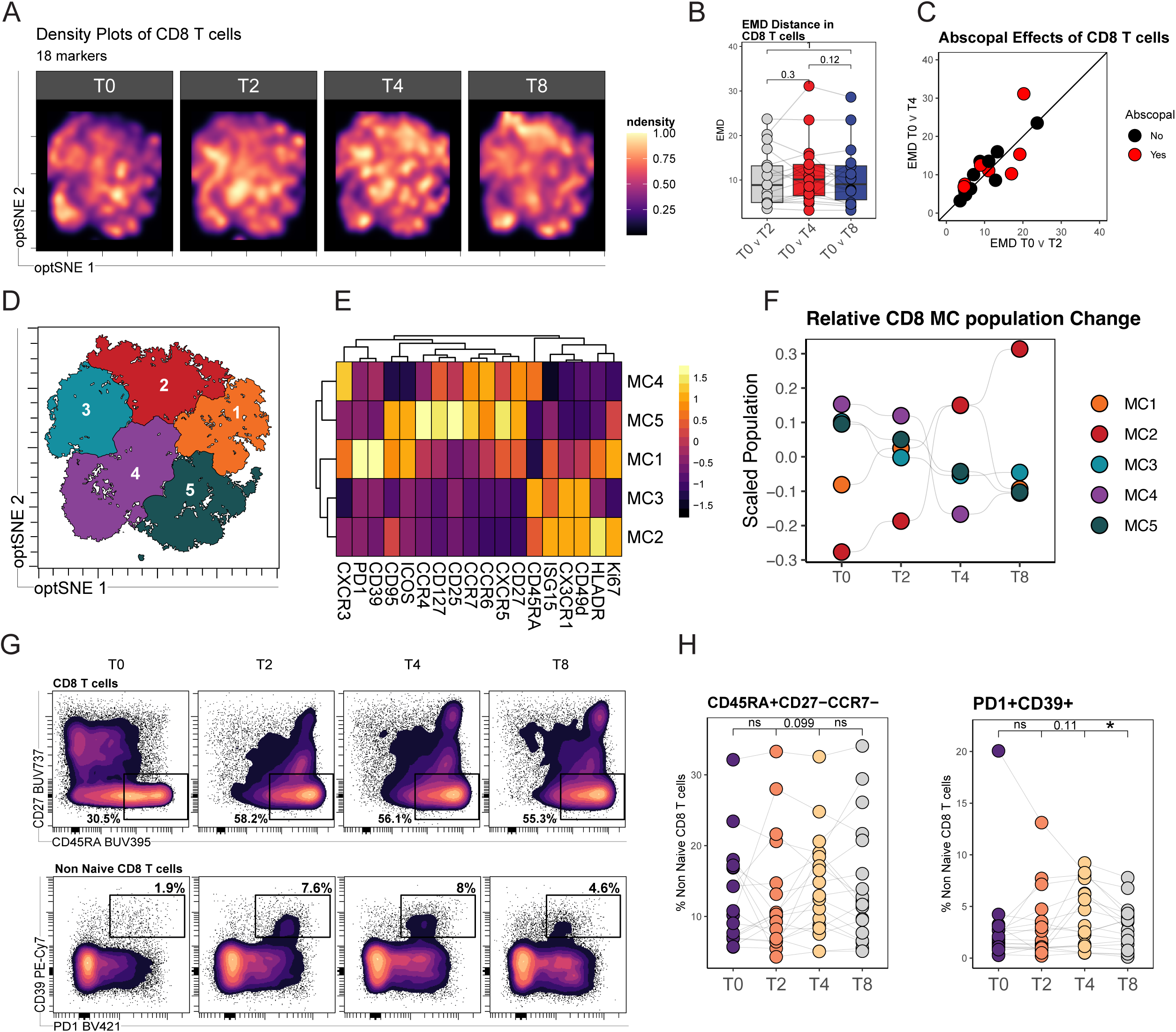
(A) Density plots of 500K CD8 T cells equally sampled by timepoint. (B) Earth mover distance between patient optSNE projections of CD8 T cells at indicated timepoints. Significance was determined using a paired Wilcox (C) Scatter plot of EMD distance between T0 – T2 (x axis) and T0 – T4 (y axis) with black line representing a slope of 1 and color representing Abscopal response. (D) FlowSOM clustering of 500K CD8 T cells broken into 5 meta clusters. (E) Heatmap highlighting the gMFI marker expression for each meta cluster (F) Summary bump plot of scaled MC populations over time (G) Representative flow plots of EMRA (CD45RA-CD27+) CD8 non naïve T cells (top row) or activated exhausted CD8 non naïve T cells (PD1+CD39+). (H) Paired analysis of patients at 4 times points measuring TEMRA or exhausted CD8 T cells. Significance was determined using a paired Wilcox test with * p<0.05.

**SFigure 5.**
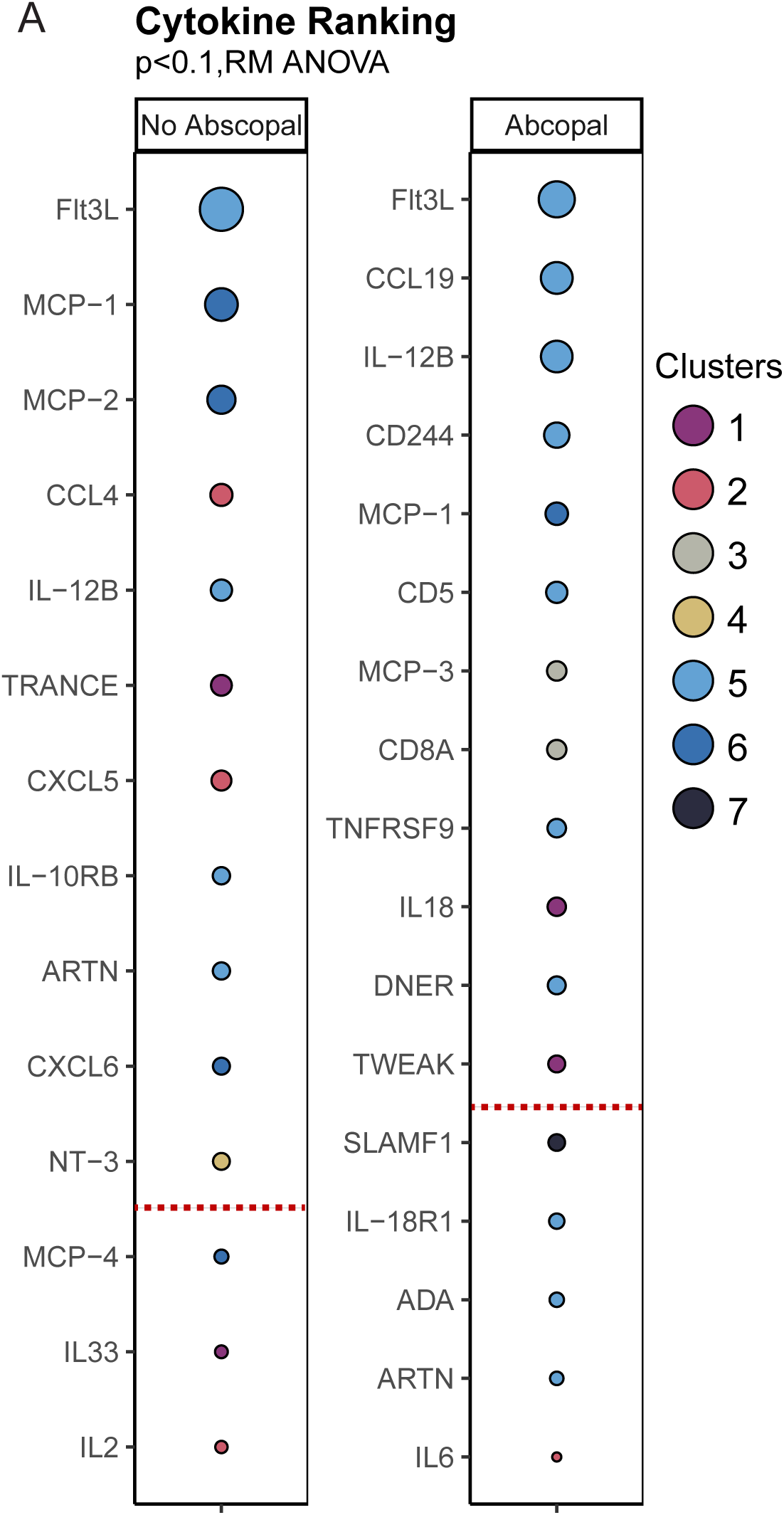
(A) Ranked list in descending order of cytokines with the have a p value <0.1 by repeated measure ANOVA. Cytokines above red lines have a p value < 0.05. Cytokines are colored by cluster group (Fig 4B) and faceted by Abscopal response.

**SFigure 6.**
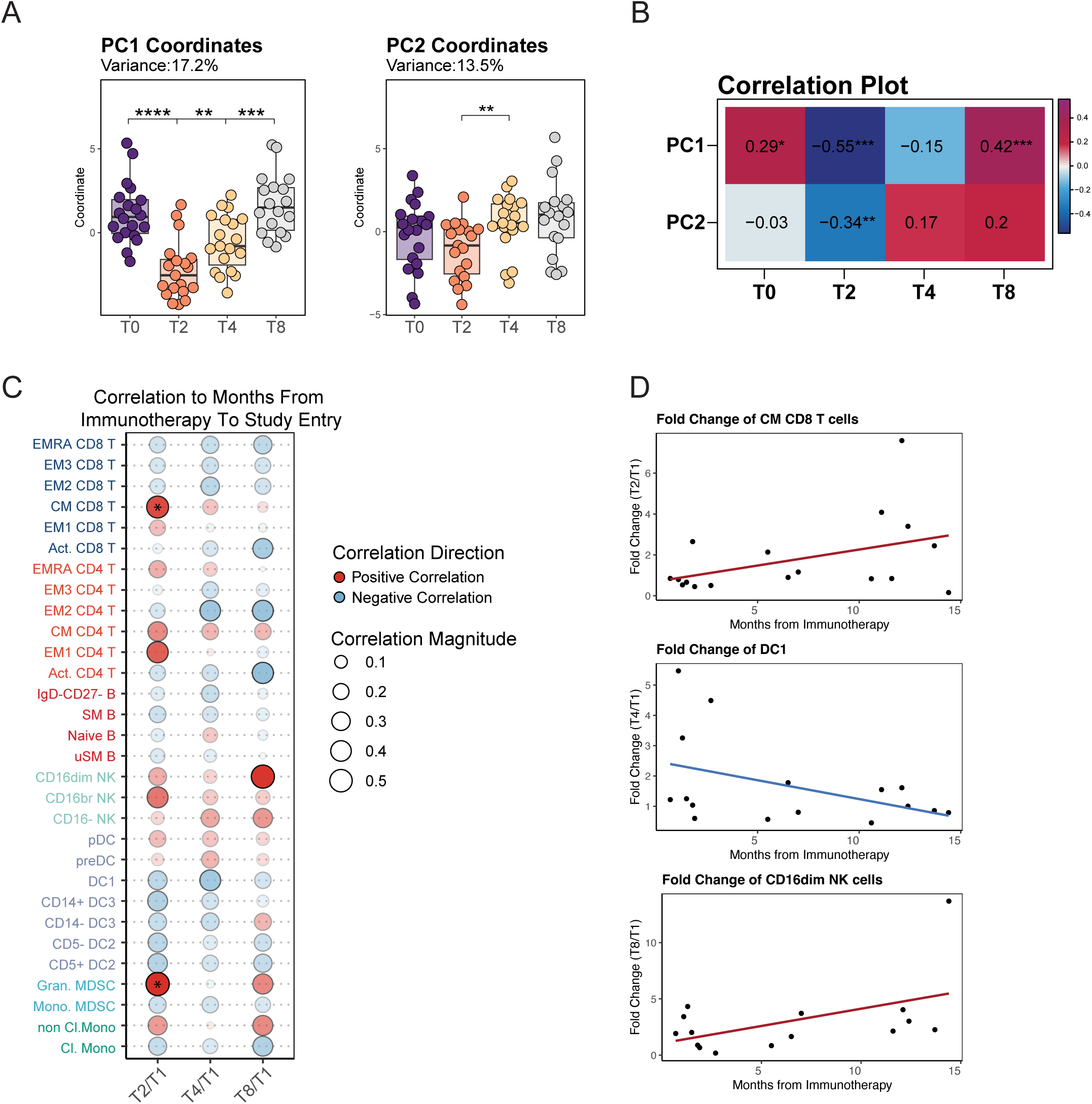
(A) PC1 (left) and PC2 coordinates (right) colored by time. Significance was determined using a Wilcox test with p<0.05,** p<=0.01, *** p<=0.001, **** p<=0.0001. (B) Correlation plot of PC1 and PC2 axis against T0, T2, T4, T8 using Pearson correlation to calculate the listed p values. (C) Fold change from baseline to T2, T4, or T8 of 31 Immune Populations to time since last immunotherapy. Pearson correlation coefficient is represented by the dot size, directionally of correlation is reflected by color, and significance marked by *,p<0.05. (D) Represented correlation of CM CD8 T cells, DC1, and CD16dim NK cells with time from last immunotherapy.

**SFigure 7.**
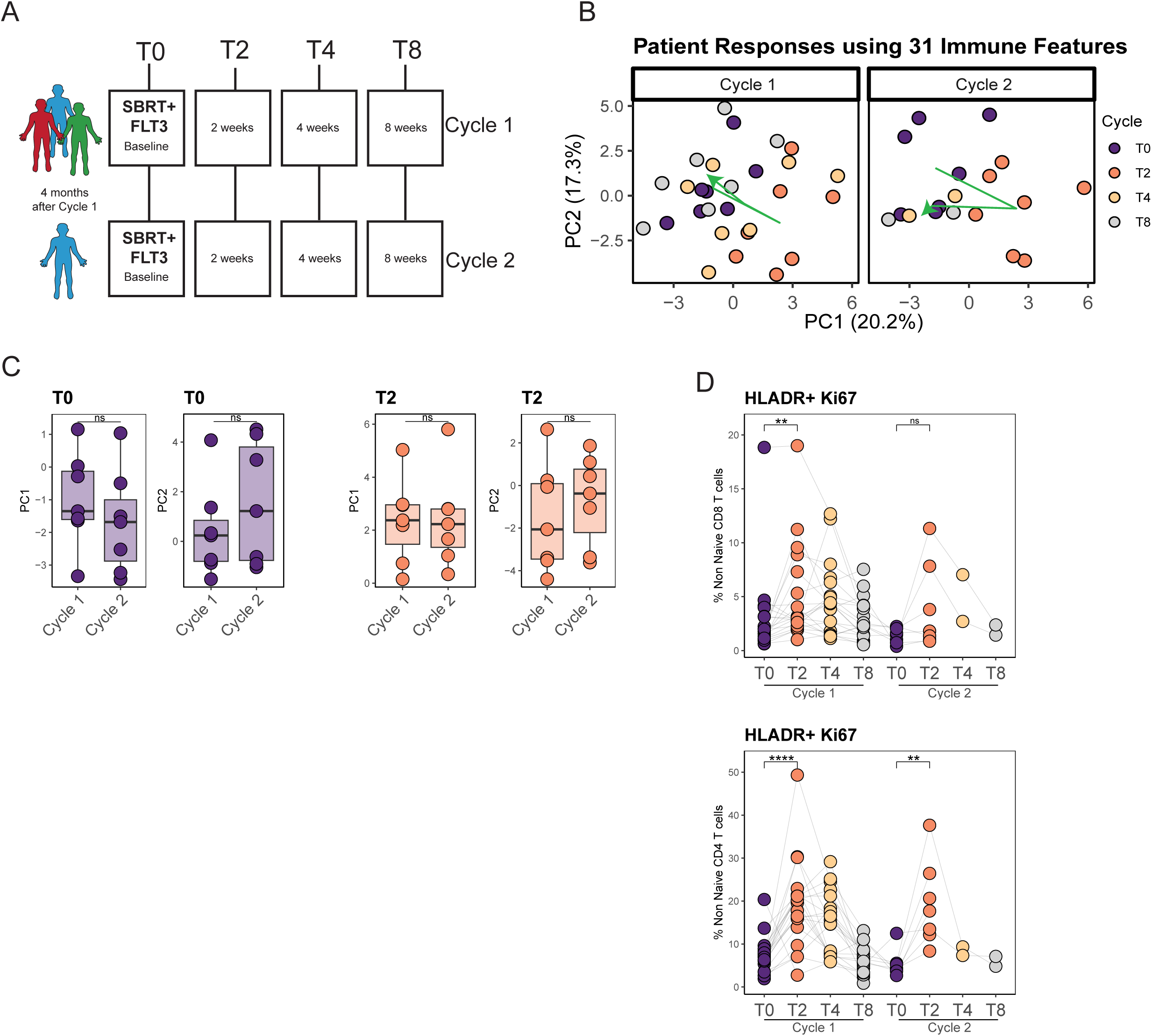
(A) Trial scheme of patients getting 2 cycles of CDX-301 and SBRT (B) Patient level PCA using aggregated flow cytometry data representing 31 unique immune populations. Each point is colored by timepoint with the green arrow representing the mean of PC1 and PC2 at each timepoint. Patients who were treated with FLT3L and SBRT in cycle 1 and cycle 2 were selected.(C) Comparison of cycle 1 and cycle 2 PC1 coordinates (left) or PC2 (right) coordinates at T0. (D) Paired analysis measuring HLADR+Ki67+ in non naïve CD8 T (top) or non naïve CD4 T (bottom) changing over two cycles of FLT3L and SBRT. Significance was determined using a paired Wilcox test with ns p>0.05, * p<0.05,** p<=0.01, *** p<=0.001, **** p<=0.0001.

**SFigure 8.**
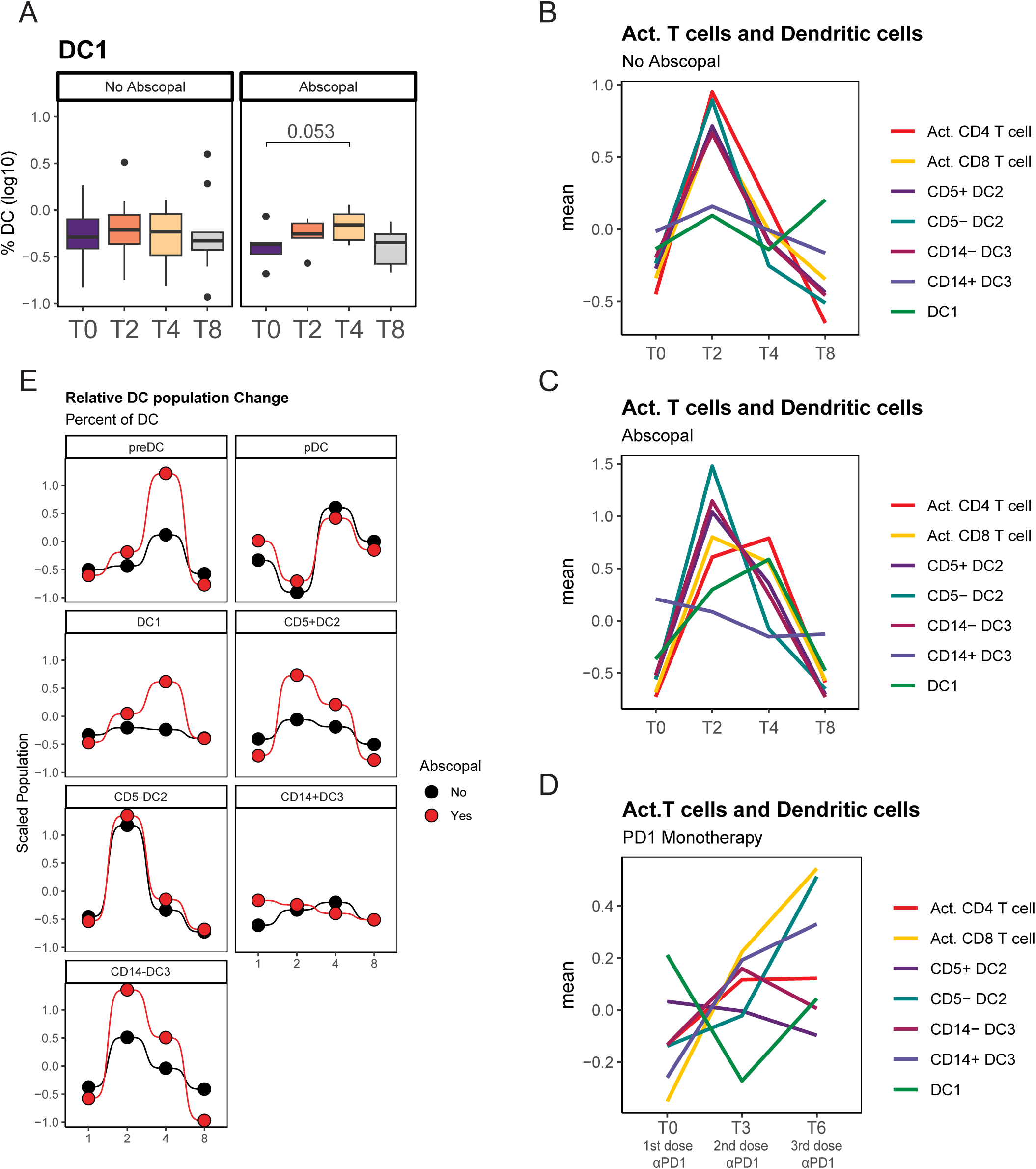
(A) Paired analysis measuring DC1 changes n HLADR+ cells faceted by Abscopal responses. Significance was determined using a paired Wilcox test. (B-D) Average of scaled populations of DC subsets or activated T cell subsets changing with time in non abscopal responders or abscopal responders (C) after FLT3L and SBRT or after (D) PD1 monotherapy. (E) Bump plot of the median scaled value of DC populations changing over time separated by DC subsets and colored by abscopal response group.

## References

1. Morabito, A. Second-line treatment for advanced NSCLC without actionable mutations: is immunotherapy the ‘panacea’ for all patients? BMC Med 16, (2018).

2. Gubens, M. A. & Wakelee, H. A. Docetaxel in the treatment of non-small cell lung carcinoma: an update and analysis. Lung Cancer: Targets and Therapy 1, 63 (2010).

3. Hanna, N. et al. Randomized phase III trial of pemetrexed versus docetaxel in patients with non-small-cell lung cancer previously treated with chemotherapy. Journal of Clinical Oncology 22, 1589–1597 (2004).

4. Giles, J. R., Globig, A.-M., Kaech, S. M. & Wherry, E. J. CD8 + T cells in the cancer-immunity cycle. doi:10.1016/j.immuni.2023.09.005.

5. Sia, J., Szmyd, R., Hau, E. & Gee, H. E. Molecular Mechanisms of Radiation-Induced Cancer Cell Death: A Primer. Front Cell Dev Biol 8, 512111 (2020).

6. Elizabeth Nelson, B., Adashek, J. J., Lin, S. H. & Subbiah, V. The abscopal effect in patients with cancer receiving immunotherapy. doi:10.1016/j.medj.2023.02.003.

7. Postow, M. A. et al. Immunologic Correlates of the Abscopal Effect in a Patient with Melanoma. New England Journal of Medicine 366, 925–931 (2012).

8. Twyman-Saint Victor, C., et al. Radiation and dual checkpoint blockade activate non-redundant immune mechanisms in cancer. Nature 2015 520:7547 520, 373–377 (2015).

9. Lynch, D. H. et al. Flt3 ligand induces tumor regression and antitumor immune responses in vivo. Nat Med 3, 625–631 (1997).

10. Braun, S. E. et al. Flt3 ligand antitumor activity in a murine breast cancer model: a comparison with granulocyte-macrophage colony-stimulating factor and a potential mechanism of action. Hum Gene Ther 10, 2141–2151 (1999).

11. Somers, K. D. et al. Orthotopic treatment model of prostate cancer and metastasis in the immunocompetent mouse: Efficacy of flt3 ligand immunotherapy. Int J Cancer 107, 773– 780 (2003).

12. Chakravarty, P. K. et al. Flt3-Ligand Administration after Radiation Therapy Prolongs Survival in a Murine Model of Metastatic Lung Cancer.

13. Fong, L. et al. Altered peptide ligand vaccination with Flt3 ligand expanded dendritic cells for tumor immunotherapy. Proc Natl Acad Sci U S A 98, 8809–8814 (2001).

14. Bhardwaj, N. et al. Flt3 ligand augments immune responses to anti-DEC-205-NY-ESO-1 vaccine through expansion of dendritic cell subsets. Nat Cancer 1, 1204–1217 (2020).

15. Huang, A. C. et al. T-cell invigoration to tumour burden ratio associated with anti-PD-1 response. Nature 545, 60 (2017).

16. Liu Yong-Jun, Kanzler, H., Soumelis, V. & Gilliet, M. Dendritic cell lineage, plasticity and cross-regulation. Nature Immunology 2001 2:7 2, 585–589 (2001).

17. Bourdely, P. et al. Transcriptional and Functional Analysis of CD1c+ Human Dendritic Cells Identifies a CD163+ Subset Priming CD8+CD103+ T Cells. Immunity 53, 335–352.e8 (2020).

18. Dutertre, C. A. et al. Single-Cell Analysis of Human Mononuclear Phagocytes Reveals Subset-Defining Markers and Identifies Circulating Inflammatory Dendritic Cells. Immunity 51, 573–589.e8 (2019).

19. Karsunky, H., Merad, M., Cozzio, A., Weissman, I. L. & Manz, M. G. Flt3 ligand regulates dendritic cell development from Flt3+ lymphoid and myeloid-committed progenitors to Flt3+ dendritic cells in vivo. J Exp Med 198, 305–313 (2003).

20. Mellman, I. & Steinman, R. M. Dendritic cells: Specialized and regulated antigen processing machines. Cell 106, 255–258 (2001).

21. Veglia, F., Sanseviero, E. & Gabrilovich, D. I. Myeloid-derived suppressor cells in the era of increasing myeloid cell diversity. Nature Reviews Immunology 2021 21:8 21, 485–498 (2021).

22. Orlova, D. Y. et al. Earth Mover’s Distance (EMD): A True Metric for Comparing Biomarker Expression Levels in Cell Populations. PLoS One 11, (2016).

23. Deaglio, S. et al. Adenosine generation catalyzed by CD39 and CD73 expressed on regulatory T cells mediates immune suppression. Journal of Experimental Medicine 204, 1257–1265 (2007).

24. Zwijnenburg, A. J. et al. Graded expression of the chemokine receptor CX3CR1 marks differentiation states of human and murine T cells and enables cross-species interpretation. Immunity 56, 1955–1974.e10 (2023).

25. Ng Tang, D., et al. Increased frequency of ICOS+ CD4 T cells as a pharmacodynamic biomarker for anti-CTLA-4 therapy. Cancer Immunol Res 1, 229–234 (2013).

26. Huang, A. C. et al. T-cell invigoration to tumour burden ratio associated with anti-PD-1 response. Nature 545, 60 (2017).

27. Kamphorst, A. O. et al. Proliferation of PD-1+ CD8 T cells in peripheral blood after PD-1-targeted therapy in lung cancer patients. Proc Natl Acad Sci U S A 114, 4993–4998 (2017).

28. Hsieh, C. S. et al. Development of TH1 CD4+ T Cells Through IL-12 Produced by Listeria-Induced Macrophages. Science (1979) 260, 547–549 (1993).

29. Parajuli, P. et al. Flt3 ligand and granulocyte-macrophage colony-stimulating factor preferentially expand and stimulate different dendritic and T-cell subsets. Exp Hematol 29, 1185–1193 (2001).

30. Mathew, D. et al. Durable Response and Improved CD8 T Cell Plasticity in Lung Cancer Patients After PD1 Blockade and JAK Inhibition. medRxiv 2022.11.05.22281973 (2022) doi:10.1101/2022.11.05.22281973.

31. Beatty, G. L. & Gladney, W. L. Immune escape mechanisms as a guide for cancer immunotherapy. Clin Cancer Res 21, 687 (2015).

32. Twyman-Saint Victor, C., et al. Radiation and Dual Checkpoint Blockade Activates Non-Redundant Immune Mechanisms in Cancer. Nature 520, 373 (2015).

33. Habets, T. H. P. M. et al. Fractionated Radiotherapy with 3 x 8 Gy Induces Systemic Anti-Tumour Responses and Abscopal Tumour Inhibition without Modulating the Humoral Anti-Tumour Response. PLoS One 11, e0159515 (2016).

34. Chakravarty, P. K. et al. Flt3-Ligand Administration after Radiation Therapy Prolongs Survival in a Murine Model of Metastatic Lung Cancer.

35. Twyman-Saint Victor, C., et al. Radiation and dual checkpoint blockade activate non-redundant immune mechanisms in cancer. Nature 2015 520:7547 520, 373–377 (2015).

36. Theelen, W. S. M. E. et al. Effect of Pembrolizumab After Stereotactic Body Radiotherapy vs Pembrolizumab Alone on Tumor Response in Patients With Advanced Non–Small Cell Lung Cancer: Results of the PEMBRO-RT Phase 2 Randomized Clinical Trial. JAMA Oncol 5, 1276 (2019).

37. Chen, M. et al. Adrenergic stress constrains the development of anti-tumor immunity and abscopal responses following local radiation. doi:10.1038/s41467-020-15676-0.

38. Bhardwaj, N. et al. Flt3 ligand augments immune responses to anti-DEC-205-NY-ESO-1 vaccine through expansion of dendritic cell subsets. Nat Cancer 1, 1204–1217 (2020).

39. Fong, L. et al. Altered peptide ligand vaccination with Flt3 ligand expanded dendritic cells for tumor immunotherapy. Proc Natl Acad Sci U S A 98, 8809–8814 (2001).

40. Dutertre, C. A. et al. Single-Cell Analysis of Human Mononuclear Phagocytes Reveals Subset-Defining Markers and Identifies Circulating Inflammatory Dendritic Cells. Immunity 51, 573–589.e8 (2019).

41. Villani, A. C. et al. Single-cell RNA-seq reveals new types of human blood dendritic cells, monocytes, and progenitors. Science (1979) 356, (2017).

42. Ferris, S. T. et al. cDC1 prime and are licensed by CD4+ T cells to induce anti-tumour immunity. Nature 584, 624–629 (2020).

43. Binnewies, M. et al. Unleashing Type-2 Dendritic Cells to Drive Protective Antitumor CD4 + T Cell Immunity In Brief. doi:10.1016/j.cell.2019.02.005.

44. Bourdely, P. et al. Transcriptional and Functional Analysis of CD1c+ Human Dendritic Cells Identifies a CD163+ Subset Priming CD8+CD103+ T Cells. Immunity 53, 335–352.e8 (2020).

45. Maier, B. et al. A conserved dendritic-cell regulatory program limits antitumour immunity. Nature 580, 257–262 (2020).

46. Demaria, S. et al. Ionizing radiation inhibition of distant untreated tumors (abscopal effect) is immune mediated. Int J Radiat Oncol Biol Phys 58, 862–870 (2004).

47. Toes, R. E. M., Schoenberger, S. P., Van Der Voort, E. I. H., Offringa, R. & Melief, C. J. M. CD40-CD40Ligand interactions and their role in cytotoxic T lymphocyte priming and anti-tumor immunity. Semin Immunol 10, 443–448 (1998).

48. Espinosa-Carrasco, G. et al. Intratumoral immune triads are required for immunotherapy-mediated elimination of solid tumors. Cancer Cell 42, 1202–1216.e8 (2024).

49. Chen, D. et al. CTLA-4 blockade induces a microglia-Th1 cell partnership that stimulates microglia phagocytosis and anti-tumor function in glioblastoma. Immunity 56, 2086–2104.e8 (2023).

50. Kruse, B. et al. CD4+ T cell-induced inflammatory cell death controls immune-evasive tumours. Nature 618, 1033–1040 (2023).

51. Astier, A. L. et al. RNA Interference Screen in Primary Human T Cells Reveals FLT3 as a Modulator of IL-10 Levels. The Journal of Immunology 184, 685–693 (2010).

52. Bourdely, P. et al. Transcriptional and Functional Analysis of CD1c+ Human Dendritic Cells Identifies a CD163+ Subset Priming CD8+CD103+ T Cells. Immunity 53, 335–352.e8 (2020).

53. Villani, A. C. et al. Single-cell RNA-seq reveals new types of human blood dendritic cells, monocytes, and progenitors. Science (1979) 356, (2017).

